# Incident benzodiazepine and Z-drug use and subsequent risk of alcohol- and drug-related problems: a nationwide matched cohort study with co-twin comparison

**DOI:** 10.1101/2024.11.27.24318091

**Authors:** Xinchen Wang, Zheng Chang, Yasmina Molero, Kayoko Isomura, Lorena Fernández de la Cruz, Paul Lichtenstein, Ralf Kuja-Halkola, Brian M D’Onofrio, Patrick D Quinn, Henrik Larsson, Isabell Brikell, Clara Hellner, Jan Hasselström, Nitya Jayaram-Lindström, David Mataix-Cols, Anna Sidorchuk

## Abstract

**Background:** Despite considerable interest in the consequences of benzodiazepine and benzodiazepine-related Z-drug (BZDR) use, little is known about whether and how initiation of BZDR treatment contributes to the development of alcohol- and drug-related problems.

**Aim:** To examine the association of incident BZDR dispensing with subsequent development of broadly defined alcohol- and drug-related problems.

**Methods:** This nationwide register-based study included demographically matched and co-twin control cohorts. Among all Swedish residents aged ≥10 years and BZDR-naïve by 2007, 960,430 BZDR-recipients with incident dispensation in 2007-2019 and without any recorded pre-existing substance-related conditions were identified and matched (1:1) to nonrecipients from the general population. Twin BZDR-recipients (n=12,048) were linked to 12,579 unexposed co-twins. Outcomes included alcohol and drug use disorders, poisoning, deaths, and related suspected criminal offences. Flexible parametric survival models estimated outcome risks across up to 14 years of follow-up.

**Results:** In the demographically matched cohort (60% women, median age at BZDR initiation 51 years), incidence rates in BZDR-recipients and nonrecipients (per 1000 person-years) were 5.60 vs 2.79 for alcohol-related and 4.15 vs 1.23 for drug-related problems, respectively. In fully-adjusted models, relative risks were increased for alcohol- and drug-related problems (adjusted hazard ratio [95% confidence interval]: 1.56 [1.53-1.59] and 2.11 [2.05-2.17], respectively). The risks persisted within the co-twin comparison, different follow-ups, and all additional and sensitivity analyses.

**Conclusions:** BZDR initiation was associated with a small but robust increase in absolute and relative risks of developing alcohol- and drug-related problems. The findings contribute to evidence base for making decisions on BZDR treatment initiation.

## Introduction

Benzodiazepines are effective in alleviating the symptoms of anxiety and insomnia and are recognized as a treatment option for managing acute alcohol withdrawal.^1^ Benzodiazepine-related Z-drugs, structurally similar to benzodiazepines, are widely used for treating insomnia.^2^ Benzodiazepines and Z-drugs (henceforth ‘BZDR’, if mentioned together) are generally considered safe and effective when used for a short period of time, as recommended by clinical guidelines.^3^ However, the risk of dependence and tolerance increases with prolonged use. Although prescription BZDR are rarely reported as the primary substance of abuse, they are often involved in polysubstance use,^4–6^ particularly with opioids and alcohol.^7–9^ Furthermore, the concurrent use of BZDR with other substances has been widely acknowledged as a contributor to deaths due to overdose.^10,11^ Nevertheless, studies on concurrent use of BZDR with other substances are mainly based on individuals with pre-existing substance use disorders.^8–12^ Yet, it is unclear to what extent BZDR use per se might contribute to the occurrence of other substance use in future.

Understanding how indicated use of BZDR, particularly among new recipients, is associated with subsequent risk of substance use disorders and related problems could be essential for preventing the development of such conditions among patients who might benefit from BZDR treatment. Some research suggests that BZDR use among individuals undergoing opioid treatment could be a risk factor for transitioning to chronic and high-dose opioid use,^13–15^ whereas those studies focus specifically on changes in prescribed opioid use in the context of pain management. Less is known about BZDR prescriptions and the risk of other substance-related disorders. One retrospective cohort study^16^ reported that incident BZDR prescription was associated with increased risks of alcohol and drug use disorders, while the findings varied between different substance-related outcomes in two other studies.^17,18^ Although informative about potential associations between BZDR and substance-related problems, those studies focused on self-reported lifetime BZDR use^17,18^ and specific populations (i.e., patients with chronic non-cancer pain,^13,15^ postoperative pain^14^ or anxiety^16^). They could also be limited by potential residual confounding from genetic and environmental factors shared within families, which are important to address given a familial transmission of substance dependence^19^ and possible familial co-aggregation between substance use and the disorders for which BZDR is commonly prescribed (e.g., anxiety disorders, depression disorders, and insomnia).^20,21^

In the present study, we examined the association between BZDR prescription initiation and the subsequent development of broadly defined alcohol- and drug-related problems that resulted in treatment-seeking, deaths or suspected criminal offences, using nationwide Swedish registers. We used a demographically matched cohort and a co-twin control cohort to mitigate various confounding factors, including shared familial confounding.

## Methods

The study was approved by the Swedish Ethical Review Authority (reference number 2020-06540). The requirement for informed consent was waived because the study is register-based and all individual data were de-identified.

### Study population

Using the unique identification numbers assigned to all Swedish residents,^22^ we linked several nationwide registers with data available through December 31, 2020. The study population included individuals who 1) were 10 to 75 years old and had no dispensation records of any BZDRs in the Prescribed Drug Register^23^ by January 1, 2007; 2) had no lifetime epilepsy diagnosis, according to the National Patient Register^24^; and 3) had identifiable biological parents in the Multi-Generation Register.^25^ This yielded a study population of 6,195,245 individuals (**Figure 1**). The Prescribed Drug Register incorporates records of the prescription medications (using Anatomical Therapeutic Chemical [ATC] Classification System codes) dispensed across all pharmacies in Sweden since July 1, 2005. Therefore, all study participants had at least 1.5-year BZDR dispensation-free washout. The National Patient Register holds inpatient and specialist outpatient care data since 1973 and 2001, respectively, with the diagnoses recorded by the Swedish version of the International Classification of Diseases (ICD). The Multi-Generation Register records kinship data for all individuals born in Sweden from 1932 or resided in the country since 1961. **Supplementary Note 1** overviews all registers used in the study.

**Figure 1.**
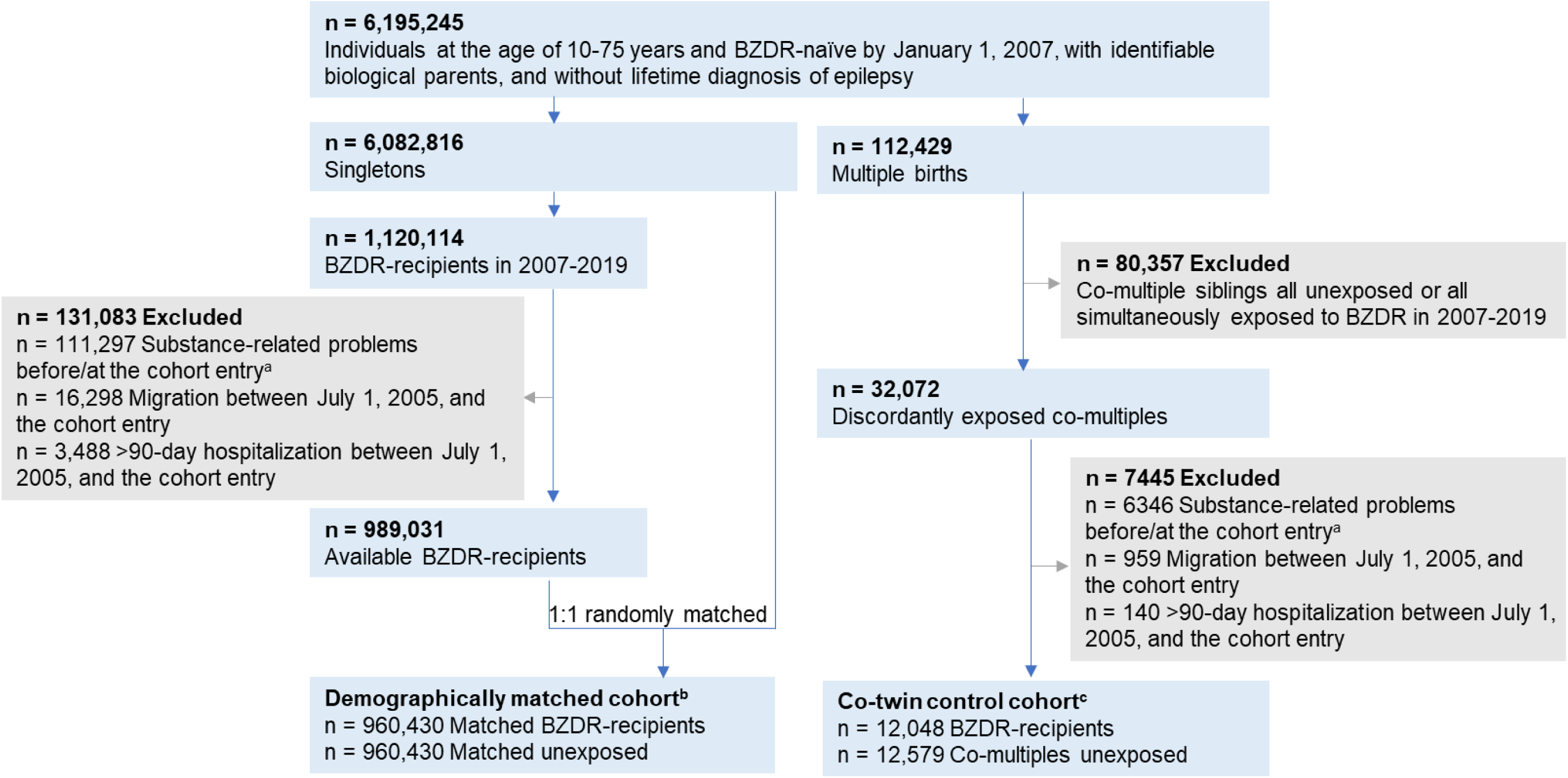
Study design overview: construction of demographically matched cohort and co-twin control cohort. ^a^ ‘Cohort entry’ refers to the date when the first prescription of any benzodiazepines or related Z-drugs (BZDR) was dispensed to the BZDR-recipient. The same date is assigned as the cohort entry to matched unexposed individual (in the demographically matched cohort) and to unexposed co-multiples (in the co-twin control cohort) ^b^ BZDR-recipients were 1:1 randomly matched by birth year and month, sex, and country of residence at the cohort entry with those who did not have any BZDR dispensation before or at the cohort entry date. To unexposed comparators, the same exclusion criteria were applied as for BZDR-recipients (i.e., no substance-related problems before or at the cohort entry, and no migration or >90-day hospitalization between July 1, 2005, and the cohort entry) ^c^ Co-multiples (of which 97.1% were twin pairs) were identified from 12,047 discordantly exposed families

### Study design and exposure to incident BZDR use

We created a demographically matched cohort using the singleton-born individuals from the study population (n=6,082,816) (**Figure 1**). Among them, we selected incident BZDR-recipients as individuals with a first BZDR dispensation recorded in the Prescribed Drug Register in 2007-2019 (n=1,120,114) with ATC-codes for benzodiazepine derivatives in anxiolytics (N05BA), hypnotics/sedatives (N05CD), antiepileptics (N03AE), and Z-drugs (N05CF) (**Supplementary Table S1**). The date of the first BZDR dispensation denoted the cohort entry for each BZDR-recipient and their matched comparator. We excluded BZDR-recipients if they: 1) were hospitalized for over 90 days between the start of washout period (July 1, 2005) and the cohort entry (this was done to further clarify the exposure status since the Prescribed Drug Register does not cover in-hospital medication); 2) emigrated from or immigrated to Sweden during that period; or 3) had records of any alcohol- or drug-related problems (**Supplementary Table S2**) before or at the cohort (to ensure incident outcome status and mitigate reverse causality). The remaining 989,301 incident BZDR-recipients were eligible for matching to individuals who had no BZDR dispensation record by the cohort entry and were not excluded due to abovementioned criteria. Matching without replacement was performed 1:1 on birth year and month, sex, and county of residence at the cohort entry, yielding 960,430 pairs of incident BZDR-recipients and their matched unexposed comparators in the demographically matched cohort.

To further control for shared genetic and environmental confounders,^26^ we constructed a co-twin control cohort using data from the Multi-Generation Register. We identified 112,429 multiple-birth individuals and assigned a family identifier to each family (**Figure 1**). Among them, 32,072 co-multiples were discordant by exposure to BZDR, i.e., at least one co-multiple in the family had incident BZDR dispensation in 2007-2019 (the date of the first BZDR dispensation was the cohort entry), and the other co-multiple(s) had no BZDR dispensations by the cohort entry (the same date was set as the cohort entry for unexposed co-twin). After applying the same exclusion criteria as above, we obtained 12,048 BZDR-recipients and their 12,579 unexposed co-multiples (97.1% were twin-pairs) from 12,047 families to form the co-twin control cohort.

### Alcohol-related problems and drug-related problems

We defined incident alcohol- and drug-related problems as two distinct outcomes using clinical, pharmacological, criminal, and cause of death data.^27^ **Supplementary Table S2** provides details on outcomes data collection. For “any alcohol-related problems”, we gathered data on alcohol use disorders and poisoning by alcohol diagnosed in inpatient or specialized outpatient care (main or additional diagnoses) from the National Patient Register, and data on death causes related to alcohol (underlying or secondary causes) from the Cause of Death Register.^28^ We also retrieved records of dispensed medication for alcohol dependence from the Prescribed Drug Register and alcohol-related suspected criminal offences from the Register of People Suspected of Offences.^29^ Similarly, we collected data on “any drug-related problems” including diagnoses of drug use disorders, poisoning by drugs, death causes related to drug use, dispensed medication for opioid use disorder, and drug-related suspected criminal offences, using the same registers as above.^23,24,28,29^

### Covariates

For each member of the demographically matched cohort, we collected data on potential confounders in addition to the matching variables (i.e., birth year and month, sex, and county of residence). Disposable family income at the cohort entry (in tertiles) was retrieved from the Longitudinal Integration Database for Health Insurance and Labour Market Studies^30^ as a proxy for socioeconomic status. Calendar year of the cohort entry (2007-2009, 2010-2012, 2013-2015, 2016-2019) was included to account for a cohort effect. Given the complex interplay between various disorders and substance use, we created two binary variables based on diagnoses recorded with ICD-10 codes in the National Patient Register between 1997 and the cohort entry: 1) history of psychiatric conditions (other than the outcomes-related) and 2) history of chronic pain-related conditions^31^ (**Supplementary Table S3**). From the Prescribed Drug Register, we retrieved data on concomitant dispensations (within 3 months before the cohort entry) of other psychotropic, antiepileptic, and analgesic medications (**Supplementary Table S1**) in a single binary variable. We also collected maternal and paternal lifetime histories of alcohol- and/or drug-related problems, using the same broad definitions as for the outcomes (**Supplementary Table S2**), and created one binary variable per parent indicating “substance-related problems”. In the co-twin control cohort, the same covariates were used, except birth year and month and parental lifetime substance-related problems.

### Statistical analyses

We fitted flexible parametric survival models^32^ to estimate average and time-varying hazard ratios (HRs) and 95% confidence intervals (CIs) for the association of incident BZDR use with the risk of developing alcohol-related problems and, separately, drug-related problems. Time since the cohort entry was the underlying time scale. In both the demographically matched and co-twin control cohorts, individuals were followed from the cohort entry until the date of the incident outcome, emigration, death by non-outcome, change in exposure status among unexposed (if they were dispensed BZDRs after the cohort entry) or December 31, 2020, whichever came first.

We first ran a minimally-adjusted model. In the demographically matched cohort this model was conditioned on the matching variables, and in the co-twin control cohort it was conditioned on the family identifier and adjusted for sex. Next, in both cohorts, we applied an adjusted model by additionally controlling for all covariates described above for each cohort. Finally, in a fully-adjusted model, we accounted for a possible co-occurrence of alcohol- and drug-related problems by additionally controlling for drug-related problems in the analysis of alcohol-related outcomes, if occurred during the follow-up, and vice versa. Moreover, we estimated time-varying HRs, standardized cumulative incidence^33^ and cumulative incidence difference throughout the follow-up and within 1 year, 3 years, 5 years, and 10 years of follow-up. All time-varying models were fully-adjusted, with the baseline hazard of 5 degrees of freedom and the effect of BZDR exposure of 3 degrees of freedom.

We conducted three additional analyses using the demographically matched cohort only (due to lack of power in the co-twin control cohort). **Supplementary Note 2** describes the methods for each additional analysis. First, we categorised any alcohol-related problems and, separately, any drug-related problems by type of incident outcome event into: 1) non-fatal disorders, 2) non-fatal poisoning, 3) deaths, and 4) suspected criminal offences. Specifically for drug-related problems, we also assessed sedatives/hypnotics use disorders and related deaths as an additional type of outcome event of interest. Main analyses were repeated for each type of outcome events using the other types as competing risk. Second, we performed subgroup analyses by participant characteristics and type of initial medication (benzodiazepines or Z-drugs). Third, we calculated the cumulative dosage of BZDR by summing up the defined daily doses (DDD) dispensed during the first year after BZDR initiation, and then assessed the outcome risk by the quartile categories of cumulative BZDR dosage with the follow-up starting from the second year. Lastly, we performed a sensitivity analysis using the demographically matched cohort, where we repeated the analyses for drug-related problems after excluding dispensed medication for opioid use disorders from the outcome definition to avoid possible misclassification for medication used for pain management.^7,27^

Data management and analyses were performed in SAS version 9.4 (SAS Institute Inc.) and Stata 18.0 (StataCorp LP), respectively. All tests employed two-tailed significance set at p<0.05.

## Results

### Description of the cohorts

In the demographically matched cohort of 960,430 incident BZDR-recipients and 960,430 unexposed comparators, 60.0% were women, and the median age at the cohort entry was 51 years (interquartile range [IQR]: 37-65). Compared with unexposed individuals, higher proportions of BZDR-recipients had any psychiatric conditions (20.3% vs 8.5%), chronic pain conditions (44.6% vs 34.6%), co-medications (48.1% vs 9.9%), and maternal (3.9% vs 2.8%) or paternal (8.9% vs 7.0%) substance use-related problems (**Supplementary Table S4**).

In the co-twin control cohort of 12,048 incident BZDR-recipients and 12,579 unexposed co-twins, 61.1% and 52.0% were women, respectively, and the median age at the cohort entry was 50 years (IQR: 34-63). Compared to their unexposed co-twins, BZDR-recipients more often had psychiatric conditions (20.4% vs 8.8%), chronic pain conditions (42.1% vs 32.9%), and co-medication (49.3% vs 10.6%). Maternal and paternal substance-related problems were present in over 3% and over 7% of all co-twins, respectively (**Supplementary Table S4**).

### Incident BZDR use and alcohol-related problems

In the demographically matched cohort, we identified 40,475 incident events of any alcohol-related problems among 960,430 BZDR-recipients and 18,581 events in 960,430 matched comparators (crude incidence rates [IR] 5.60 vs 2.79 per 1000 person-years) (**Table 1**). This corresponded to a minimally-adjusted HR of 2.07 (95%CI 2.03-2.10), which attenuated in the fully-adjusted model (adjusted HR [aHR] 1.56; 95%CI 1.53-1.59). The standardized cumulative incidence of outcome in BZDR-recipients was small but constantly higher than that in unexposed individuals (e.g., at 1-year follow-up: 0.78% vs 0.44%, risk difference of 0.35% [95%CI 0.33%-0.37%]; at 10-year follow-up: 4.19% vs 2.51%, risk difference of 1.68% [95%CI 1.62%-1.74%]) (**Table 2**, **Figure 2**). The relative risk also remained increased throughout the whole study period, with aHRs (95%CIs) varying from 1.50 (1.45-1.54) at 1-year follow-up to 1.69 (1.62-1.75) at 10-year follow-up (**Table 2**, **Figure 3**). In the co-twin control cohort, we identified 438 and 170 events of alcohol-related problems among 12,048 BZDR-recipients and 12,579 unexposed co-twins, respectively (crude IR 4.75 vs 1.89 per 1000 person-years). The increased relative risk persisted in all models, including the fully-adjusted model (aHR 2.15, 95%CI 1.76-2.61) (**Table 1**). The time-varying patterns of absolute and relative risks were similar to those of the demographically matched cohort (**Table 2**, **Figures 2** and **3**).

**Figure 2.**
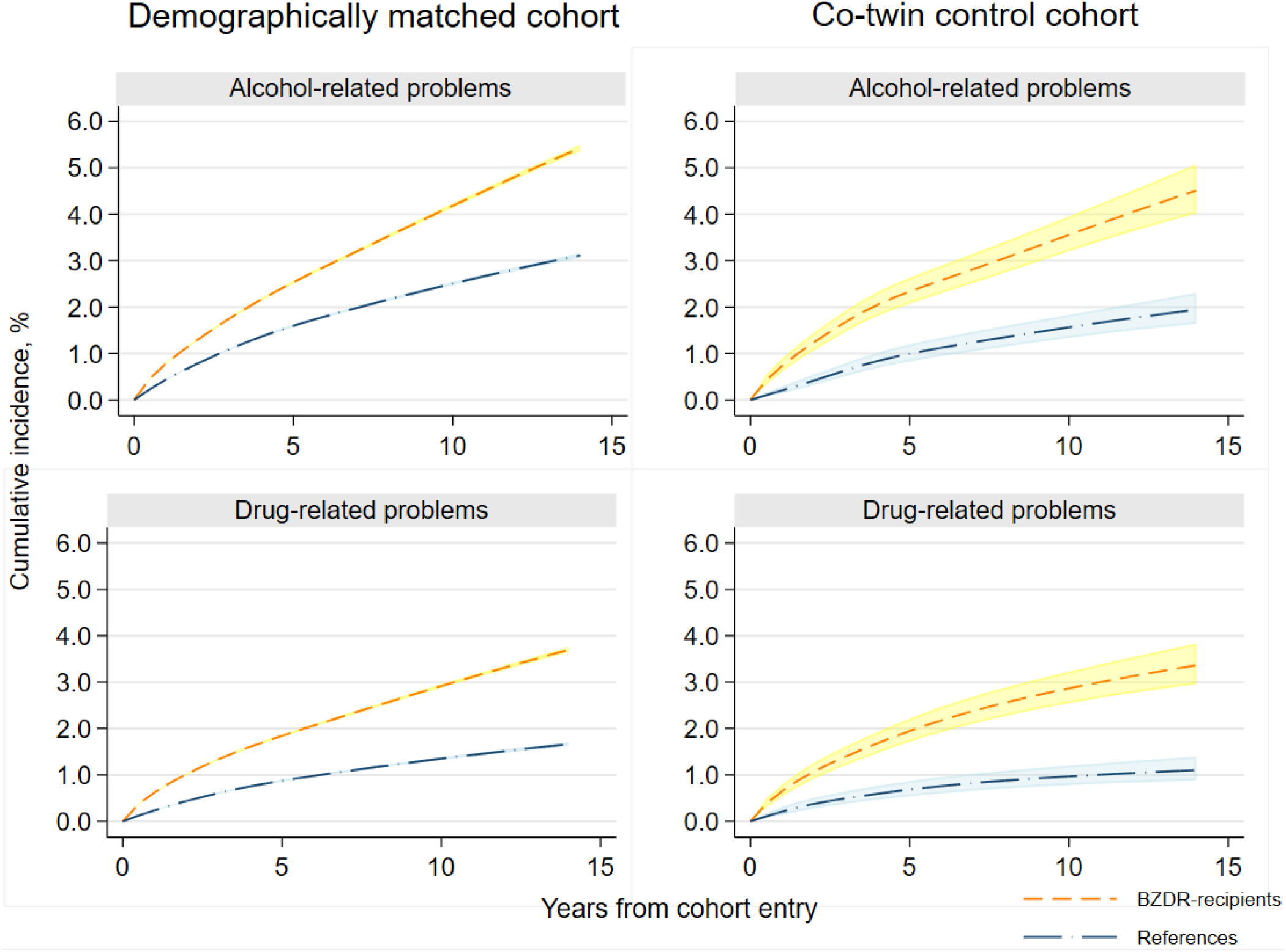
Standardized cumulative incidence and 95% CI of any alcohol-related and drug-related problems as a function of time since the cohort entry among BZDR-recipients and their matched comparators (references) in the demographically matched and co-twin control cohorts *Note*: Cumulative incidence measures are standardised, i.e., controlled for, the covariates which were included in fully-adjusted model for the corresponding cohort, and estimated from the flexible parametric model. Shadowed areas denote 95% CI. *Abbreviations*: BZDR, benzodiazepine or related Z-drugs; CI, confidence intervals.

**Figure 3.**
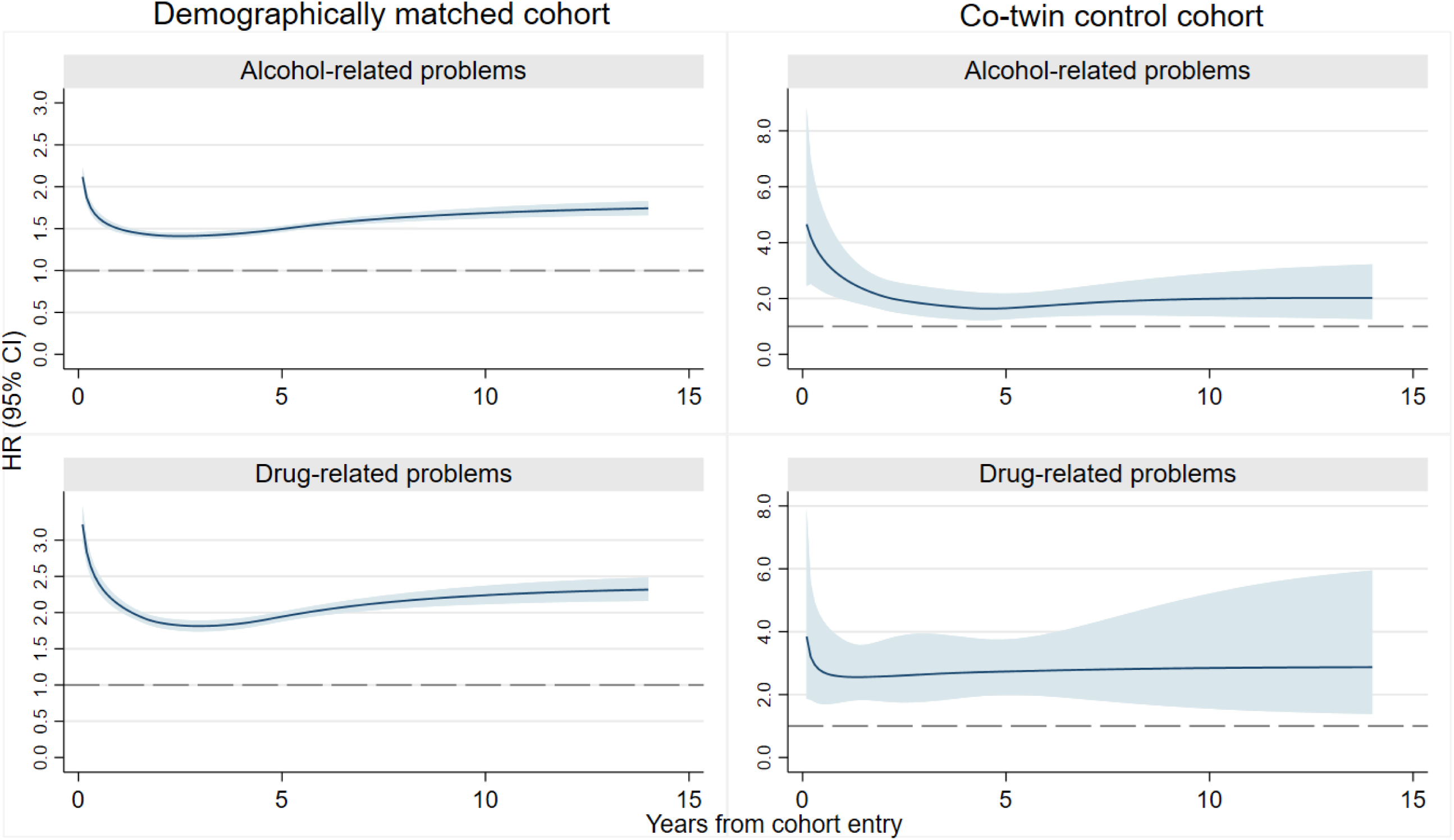
The risk of any alcohol-related and drug-related problems as a function of time since the cohort entry among BZDR-recipients and their matched comparators (references) in the demographically matched and co-twin control cohorts *Note*: All reported HRs represent the results of fully-adjusted model for the corresponding cohort. Shadowed areas denote 95% CI. *Abbreviation*: BZDR, benzodiazepine or related Z-drugs; CI, confidence intervals; HR, hazard ratio.

**Table 1.** Associations of incident BZDR use with the subsequent risk of any alcohol-related problems and any drug-related problems in 960,430 BZDR-recipients and 960,430 matched comparators in the demographically matched cohort, and among 12,048 BZDR-recipients and 12,579 unexposed co-twins in the co-twin control cohort.

|  | No. of events, n (%) |  | Crude incidence rate<br>(95% CIs), per 1000 person-years |  | Minimally<br>adjusted model<sup>ab</sup> | Adjusted<br>model<sup>cd</sup> | Fully-adjusted<br>model<sup>e</sup> |
| --- | --- | --- | --- | --- | --- | --- | --- |
|  | BZDR-recipients | Unexposed<br>individuals | BZDR-recipients | Unexposed<br>individuals | HR<br>(95% CI) | HR<br>(95% CI) | HR<br>(95% CI) |
| **Demographically matched cohort** |  |  |  |  |  |  |  |
| Any alcohol-related problems | 40,475 (4.21) | 18,581 (1.93) | 5.60 (5.55-5.66) | 2.79 (2.75-2.83) | 2.07 (2.03-2.10) | 1.60 (1.57-1.63) | 1.56 (1.53-1.59) |
| Any drug-related problems | 30,228 (3.15) | 8227 (0.86) | 4.15 (4.10-4.20) | 1.23 (1.20-1.25) | 3.55 (3.46-3.63) | 2.17 (2.11-2.23) | 2.11 (2.05-2.17) |
| **Co-twin control cohort** |  |  |  |  |  |  |  |
| Any alcohol-related problems | 438 (3.64) | 170 (1.35) | 4.75 (4.31-5.20) | 1.89 (1.61-2.18) | 2.75 (2.30-3.29) | 2.21 (1.81-2.69) | 2.15 (1.76-2.61) |
| Any drug-related problems | 360 (2.99) | 90 (0.72) | 3.88 (3.48-4.28) | 1.00 (0.79-1.20) | 4.22 (3.35-5.31) | 2.87 (2.23-3.71) | 2.78 (2.15-3.59) |

<sup>a</sup> In the demographically matched cohort: conditioned on matching variables (birth year and month, sex, country of residence at the cohort entry)
<sup>b</sup> In the co-twin control cohort: conditioned on family identifier and adjusted on sex of each co-twin
<sup>c</sup> In the demographically matched cohort: is additionally adjusted for calendar year at the cohort entry, disposable family income, history of any psychiatric conditions, chronic pain conditions, concomitant dispensations of other psychotropic, antiepileptic, and analgesic medications, if dispensed within 3 months before the cohort entry, and history of maternal and paternal substance-related problems
<sup>d</sup> In the co-twin control cohort: additionally adjusted for the same covariates as the demographically matched cohort, excluding birth year and month, and maternal and paternal substance-related problems
<sup>e</sup> In both cohorts: additionally adjusted for the events of the ‘other’ outcome if it was recorded during the study follow-up (i.e., controlling for drug-related problems in the analysis of alcohol-related problems, if occurred during the follow-up, and vice versa)
*Abbreviation:* BZDR, benzodiazepines and related Z-drugs; CI, confidence intervals; HR, hazard ratio

**Table 2.**
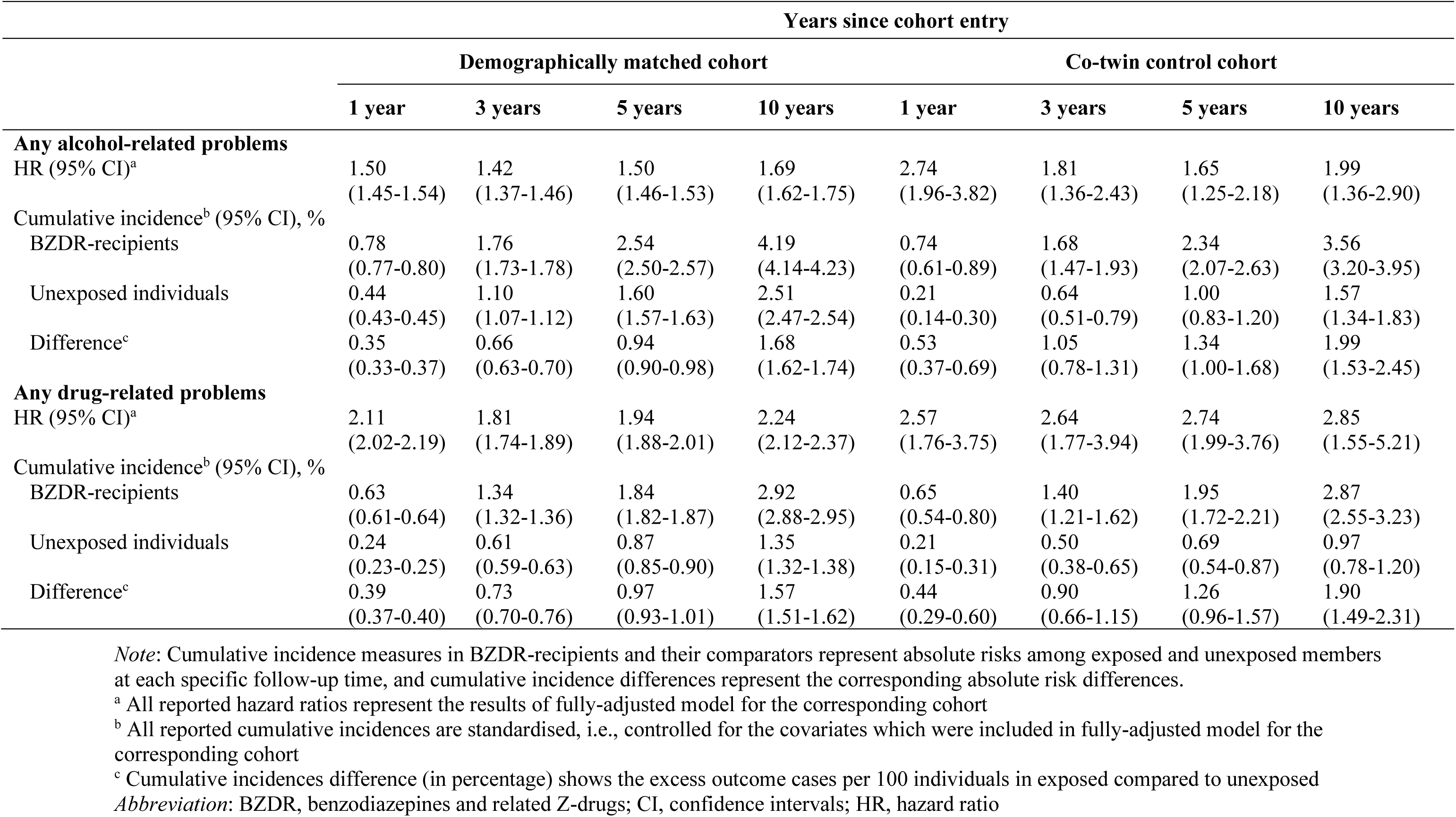
Associations of incident BZDR use with the risk of any alcohol-related problems and any drug-related problems among the members of the demographically matched cohort and the co-twin control cohort at different follow-up periods.

In the first additional analyses, BZDR use was associated with increased risks of all specific types of alcohol-related outcomes, regardless of adjustments (**Supplementary Table S5**). Within each follow-up period, the standardized cumulative incidence of the specific outcome events was fairly low (e.g., at 10-year follow-up, among the exposed it varied from 0.07% for alcohol poisoning to 3.36% for alcohol use disorders) (**Supplementary Table S6, Supplementary Figure S1**), while relative risks for most of the specific events remained increased (**Supplementary Table S6, Supplementary Figure S2**). Next, in the subgroup analysis, the associations between BZDR use and any alcohol-related problems persisted in all subgroups, with higher aHRs among women, individuals aged 10-29 years at BZDR initiation, those entering the cohort in 2016-2019, individuals without a history of diagnosed psychiatric conditions, chronic pain conditions or co-medication records, and persons who were initially dispensed benzodiazepines (**Supplementary Table S7**). Moreover, we observed a dose-response association between the cumulative dosage of BZDR use in the first year after BZDR initiation and increased relative risks of alcohol-related problems occurred from the second year after initiation (**Supplementary Table S8**).

### Incident BZDR use and drug-related problems

In the demographically matched cohort, there were 30,222 incident events of drug-related problems in BZDR-recipients and 8227 events in their matched comparators (crude IR 4.15 vs 1.23 per 1000 person-years) (**Table 1**). An over three-fold increase in relative risk was observed in the minimally-adjusted model (HR 3.55, 95%CI 3.46-3.63), which attenuated in the fully-adjusted model (aHR 2.11, 95%CI 2.05-2.17). Standardized cumulative incidence in BZDR-recipients was constantly higher than in unexposed individuals (e.g., 1-year follow-up: 0.63% vs 0.24%, risk difference of 0.39% [95%CI 0.37%-0.40%]; 10-year follow-up: 2.92% vs 1.35%, risk difference of 1.57% [95%CI 1.51%-1.62%]) (**Table 2**, **Figure 2**). The relative risk remained elevated over time, reaching an aHR of 2.24 (95%CI 2.12-2.37) at the 10-year follow-up (**Table 2**, **Figure 3**). In the co-twin control cohort, there were 360 drug-related events in BZDR-recipients and 90 events in the unexposed co-twins (crude IR 3.88 vs 1.00 per 1000 person-years), with a fully-adjusted aHR of 2.78 (95%CI 2.15-3.59) (**Table 1**). The time-varying patterns of absolute and relative risks remained similar to those in the demographically matched cohort (**Table 2**, **Figures 2** and **3**).

In the additional analysis of specific types of drug-related outcome, BZDR use was associated with increased relative risks all specific outcome events in all models. The most substantial increase in risk was seen for sedatives/hypnotics use disorders and related deaths (aHR 4.10, 95%CI 3.71-4.53) (**Supplementary Table S5**). Time-varying risks remained consistently elevated for most specific types of drug-related outcomes (**Supplementary Table S6, Supplementary Figures S3** and **S4**). In the subgroup analysis, the increased risk of drug-related problems remained in all subgroups, with higher aHRs in 10-29-year-olds, with the cohort entry in 2016-2019, those without a history of diagnosed psychiatric conditions, chronic pain conditions or co-medication records, and individuals who were initially dispensed Z-drugs (**Supplementary Tables S9**). A dose-response association was observed between the cumulative BZDR dosage and the risk of any drug-related problems (**Supplementary Table S8**). Exclusion of dispensation records for opioid use disorders medication did not alter the results (**Supplementary Table S10**).

## Discussion

In this population-based co-twin controlled study of individuals aged 10 years and above and without any recorded pre-existing substance use-related conditions, incident BZDR use was associated with the increased risks of developing severe alcohol- and drug-related problems throughout the whole 14-year study period. After controlling for various potential confounders and shared familial factors, incident BZDR-recipients had 2.11-fold and 2.78-fold higher risks of subsequent alcohol- and drug-related problems, respectively, compared to nonrecipients. The increase in relative risks persisted within different follow-up periods and in all subgroup and sensitivity analyses. Also, the risks remained elevated for the specific types of outcome events, i.e., disorders, poisoning, suspected criminal offences and deaths related to alcohol and drug use. Furthermore, a dose-response relationship was observed between cumulative BZDR dosage in the first year after initiation and increased risks of future alcohol- and drug-related problems. Finally, although the estimated incidence rates of the outcomes were fairly low, these reflect severe conditions that led to hospitalizations or specialist care, suspected criminal offences, and deaths; therefore, even a small increase in absolute and relative outcome risks warrants careful consideration.

To our knowledge, this study is the first to prospectively assess the time-varying associations of incident BZDR use and a broad range of alcohol- and drug-related problems. Although direct comparisons with previous literature are difficult due to the scarcity of studies, the average hazard ratios estimated in our study were comparable to those in a retrospective cohort study where incident benzodiazepine use was associated with a 3-fold increased risk of alcohol and drug use disorders.^16^ The slightly higher risk estimate in that study could probably be explained by the restriction to patients with anxiety and the focus on chronic benzodiazepine use.^16^

Our findings hold significant clinical importance and could contribute novel insights to existing research. First, by estimating the cumulative incidence difference, we found that for every 50 BZDR-recipients, there was approximately 1 additional case of any alcohol-related problems and 1 additional case of drug-related problems 10 years after initiation. This underscores the need for clinical monitoring of substance use behaviour among BZDR-recipients. Second, our results suggested that the associations of incident BZDR use with alcohol- and drug-related problems may not be fully explained by sociodemographic and clinical characteristics, family history of substance-related problems or shared genetic and environmental factors. Third, our assessment of the outcomes extended beyond diagnosed substance use disorders and overdoses, including also alcohol- and drug-related deaths and suspected criminal offenses. Since deaths and offences do not depend on treatment-seeking behaviour, our study provides a more comprehensive evaluation of substance-related risks. Fourth, we explored the role of BZDR characteristics (type of drugs, dosage) and observed similarly strong associations with outcomes in individuals with benzodiazepines as first medication dispensed and in those with Z-drugs as initial medication. This aligns with existing evidence suggesting that Z-drugs could also contribute to an increased risk of substance misuse, particularly to drug overdose.^7^ In the dose-response analysis, we observed an increased risk of the outcomes already at the lower category of the cumulative BZDR dosage (<30 DDD during the first year of BZDR treatment). Fifth, we observed higher relative risks of both alcohol- and drug-related problems in individuals without a recorded history of psychiatric conditions, chronic pain conditions or co-medication. Although these findings cannot fully rule out confounding by indication, our results indicate that BZDR initiation may contribute to an increased risk of subsequent substance-related problems, beyond its association with underlying mental disorders which are known to elevate such risks.^9,10^

Biological studies have suggested shared pharmacological mechanisms for BZDR and some other substances.^34^ For example, the therapeutic effects of BZDR are achieved via allosteric modulation of gamma-aminobutyric acid type A (GABA_A_) receptors, which are known to modulate the activity of brain circuits involved in the behavioural effects of drugs of abuse.^35,36^ Similar to BZDR, alcohol also acts as an agonist at GABA_A_ receptors.^37^ Further, genetic research showed that the gene encoding the α2 subunit of the GABAA receptor (GABRA2) may modulate the rewarding effects of BZDRs, alcohol, cocaine, and opioids, and thus directly influence drug-seeking behaviour.^38^ Also, it has been reported that the transition from medical BZDR use towards misuse of BZDR and other prescription medications, or towards illicit drugs and alcohol use, may be explained by the motivation to seek euphoric effects and self-medication.^21,23,24^

Our results indicate the importance for healthcare practitioners and patients to consider long-term elevated risks of alcohol and drug-related problems associated with initiating BZDR treatment. This highlights the need for prescribers not only to carefully consider the risks and benefits of BZDR prescription prior to treatment initiation, but also to prioritize monitoring and screening strategies for possible changes in patients’ substance use behaviours throughout the treatment process. Furthermore, to inform better clinical practices and strategies to mitigate the possible adverse outcomes of BZDR use, one challenge will be to understand the context in which BZDR initiation may result in elevated risks of substance-related problems. This refers to a better knowledge of, for example, how various treatment properties (e.g., the role of specific BZDR drugs, duration of treatment) and time-varying social, economic, and psychological conditions impact the risk of substance use and misuse.

### Strengths and limitations

The study strengths include using population-based Swedish registers with nationwide coverage and standardized data collection that ensured the results generalizability to the national level and minimized the risk of reporting, selection, and information bias. Also, the large sample size, long follow-up time, and a variety of health and administrative data enabled us to analyse alcohol- and drug-related problems separately and perform a range of informative additional and sensitivity analyses. Moreover, we included suspected criminal offences and deaths to the study outcomes to capture substance-related problems that occur in the non-medical context or could go untreated. Furthermore, the use of the co-twin control design allowed to account for confounding due to unmeasured genetic and environmental factors shared within families.

Several limitations should be acknowledged. First, despite broad outcome definitions, the diagnoses of alcohol and drug use disorders (which constituted most outcome events) may only represent severe cases that led to hospitalization or specialist treatment, leaving milder clinical cases uncaptured. Second, it was uncertain whether and when the BZDR-recipients consumed the dispensed medication. However, prior literature^39^ suggests that BZDR are mainly used in proximity to its dispensation date, making the register records a reasonable proxy for actual BZDR consumption. Third, data on exposure were solely based on the Prescribed Drug Register. According to surveys from the Swedish Council for Information on Alcohol and Other Drugs, the use of sedatives/hypnotics (mainly BZDRs) without doctor’s prescriptions is reported by 2% of respondents aged 17-84 years^40^ and 4-5% of late adolescents.^41^ Thus, the observed associations could be biased if illegal BZDR purchases or recreational use were disproportionally distributed between the exposure categories. Fourth, we could not fully account for lifestyle factors and their changes over time. The results from the co-twin control cohort might have partly addressed this concern because by design it controls for early life environments clustered within families. However, there are still many factors that are specific to each twin/multiple birth individual. Also, when controlling for history of psychiatric and somatic conditions, we only captured conditions that led to specialist service treatment, while the corresponding information from the primary care was unavailable. Fifth, the potential confounding by contraindication should be noted since clinical guidelines advise against prescribing BZDR to individuals with past or current substance misuse.^42,43^ Our study population was initially restricted to individuals without records of prior substance-related problems; yet, we may have missed pre-existing substance use behaviours not diagnosed/recorded in the registers. Finally, results may not be generalizable to other countries due to possible differences in BZDR prescribing and identification of substance-related events.

### Conclusions

The present study suggests an overall small but robust increase in risk of subsequent development of alcohol and drug-related morbidity, mortality, and suspected criminal offences after BZDR treatment initiation. The associations were not fully explained by sociodemographic characteristics, history of psychiatric conditions, chronic pain conditions, concomitant use of other medications, parental substance-related problems, and shared familial factors. While further research is needed to uncover the mechanisms behind the observed associations, our findings underscore the need for informed decision-making in initiating and monitoring BZDR treatment.

## Data Availability

Sharing individual-level data are restricted by Swedish data protection laws, therefore the data underlying our findings cannot be made publicly accessible

## Contributions of Authors

Conceptualization and methodology: AS, DMC, and XW. Formal analysis: XW. Interpretation of data and critical revision of the manuscript for important intellectual content: all authors. Original draft preparation: XW and AS. Funding acquisition: AS. Supervision: AS, DMC, ZC, NJ-L. Final manuscript approval: all authors. All authors have read and approved the final submitted manuscript and agree to be accountable for the work.

