## Supplements for "Incident benzodiazepine and Z-drug use and subsequent risk of alcohol- and drug-related problems: a nationwide matched cohort study with co-twin comparison"

**Supplementary materials**

**Supplementary Note 1. The Swedish nationwide registers and databases used in the study**

**1)** ***The Swedish Prescribed Drug Register***^1^ encompasses data on prescribed medications dispensed across all pharmacies in Sweden since July 2005 onwards, registered using Anatomical Therapeutic Chemical (ATC) Classification System codes, along with dosage, dispensed amount, dispensation date, and prescriber’s characteristics. The register does not include treatment indication and medications administered in hospitals. In this study, from the Prescribed Drug Register we retrieved data on the initial benzodiazepines and the related Z-drugs (BZDR), and data on other psychotropic, antiepileptic, and analgesic medications, if dispensed within 3 months prior to BZDR initiation. Further, ATC-codes for medication used for alcohol dependence and opioid used disorders were retrieved as a part of the outcome definition, as well as for defining the history of alcohol- and drug-related disorders among the study cohort members, and for parental lifetime substance-related disorders.

**2)** ***The National Patient Register***^2^ captures diagnostic information from somatic and psychiatric inpatient care (covered since 1969 and 1973, respectively) and specialist outpatient care (since 2001), based on the Swedish version of the International Classification of Diseases, Eighth Revision (ICD-8) (1969-1986), ICD-9 (1987-1996), and ICD-10 (1997-onwards). The register was validated for an array of diagnoses with an overall positive predictive value of 85-95% and up to 97% for psychiatric disorders.^3-6^ In this study, data on alcohol- and drug-related disorders and poisoning were retrieved from this register as a part of the outcome measure. Similar data were used for defining the history of alcohol- and drug-related disorders among the study cohort members, and for parental history of alcohol and drug-related disorders. Further, data on diagnosed psychiatric and somatic conditions, were retrieved from this register to be used as covariates. Finally, the register data were also used to restrict the study population to individuals without the lifetime diagnosis of epilepsy and to gain information on those who were hospitalized for longer than 90 days.

**3)** ***The Cause of Death Register***^7^ includes information on all deaths of Swedish residents, occurring in Sweden or abroad, with dates and the international version of the ICD codes for underlying and contributory causes of deaths since 1952. In this study, death data and causes of death were collected from this register and used a part of the outcome definition (i.e., death due to alcohol and drug use disorders and poisoning), and death by the cause other than the outcome as a censoring event.

**4)** ***The Total Population Register***^8^ records demographic data of all Swedish inhabitants since 1968, and ***the Migration Register*** - which is part of the Total Population Register - captures migration in and out of Sweden. In this study, data on individual’s age at the first BZDR dispensation, sex, and county of residence in Sweden were collected from the Total Population Register, while information on migration was retrieved from the Migration Register to be used for the inclusion/exclusion criteria and as a censoring event.

**5)** ***The Multi-Generation Register***^9^ contains information on biological and adopted parents of all individuals who were born in Sweden from 1932 onwards or have ever been registered in the country since 1961. With the mother as informant, the father is defined as the mother’s husband at the time of birth, or the man acknowledged as the father by unmarried mothers. The register spans over five generations and contains data on 100% of mothers and 98% of fathers for those born in Sweden since 1961.^9^ In this study, the register was used it to identify and link the biological mothers and fathers to each study participant, and to identify singleton and multiple births.

**6)** ***The Longitudinal Integration Database for Health Insurance and Labour Market Studies*** (LISA, in its Swedish acronym),^10^ since 1990 provides annual socioeconomic data for all Swedish residents aged 16 years and above. In this study, information on a disposable income at the cohort entry year (or the nearest year available) was collected from the LISA register to be used as a covariate.

**7)** ***The Register of People Suspected of Offences***,^11^ since 1995 encompasses records on all individuals aged 15 years or above (the age of criminal responsibility in Sweden) who were suspected of offences after a completed investigation by police, the customs authority, or the prosecution service. The register includes data on all reported offences even if some of them are later found not to have constituted criminal offences. In this study, information on offences related to alcohol or drug use (according to the codes corresponding to the relevant Swedish legislations on alcohol- or drug-related crimes) and the dates of offences were retrieved to be used as a part of the study outcome definition.

The Prescribed Drug Register, the National Patient Register, and the Cause of Death Register are held by the Swedish National Board of Health and Welfare. The Total Population Register, the Multi Generation Register, and the LISA register are held by Statistics Sweden. The Register of People Suspected of Offences is held by the Swedish National Council for Crime Prevention.

11. The Swedish National Council for Crime Prevention. Available at [www.bra.se](http://www.bra.se)

**Supplementary Note 2. Methods used for conducting additional analyses**

Three additional analyses were conducted using the demographically matched cohort only (sample size was insufficient in the twin cohort).

1) First, we categorised any alcohol-related problems and, separately, any drug-related problems by type of incident event into: i) *alcohol use disorders*, including the diagnoses and the records of the dispensed medication for alcohol dependence (or, correspondingly, *drug use disorders*, including the diagnoses and the records of the dispensed medication for opioid use disorder), ii) *poisoning*, as the diagnoses of poisoning by alcohol (or, correspondingly, by drugs), iii) *deaths* due to alcohol use disorders or alcohol poisoning (or, correspondingly, due to drug use disorders or poisoning by drugs), and iv) *suspected* *criminal offences* related to alcohol or drug use. The exact ICD-codes, ATC-codes, and criminal offences codes are reported in **Supplementary Table S2**.

In addition to the abovementioned types of outcome events, we were interested in studying a special type of outcome event that refers to addiction to BZDR. However, due to data availability, we were only able to collect information on sedatives/hypnotics use disorders and related deaths. More precisely, we retrieved information on non-fatal and fatal cases of sedatives/hypnotics use disorders (ICD-10 code: F13.1-13.9) and on non-fatal and fatal cases of acute intoxication/poisoning by sedatives/hypnotics (ICD-10 code: F13.0) from the National Patient Register and the Cause of Death Register, respectively. No other data on sedatives/hypnotics-related problems (or specifically on poisoning by BZDR) were available. To ensure statistical power for analysis of this type of outcome event, non-fatal and fatal cases of sedatives/hypnotics use disorders, and acute intoxications were combined in one variable.

To analyse an association of incident BZDR use with the abovementioned types of outcome events, we repeated the main analyses for each type of events using the other types of events as competing risk.

2) Second, to further explore the influence of sociodemographic characteristics, history of psychiatric conditions, chronic pain conditions, other medications dispensed in proximity to the cohort entry date, and the type of the initial BZDR medication (BZD [N03AE, N05BA, or N05CD] or Z-drugs [N05CF], which was collected at the first BZDR dispensation in 2007-2019) on associations of interest, we performed subgroup analyses. The analyses were performed for any alcohol-related problems and, separately, for drug-related problems. For each study covariate, differences between subgroup HRs were estimated by introducing interaction terms to the models or using Wald tests.

3) Third, we examined the association between the dosage of BZDR and the risk of alcohol use-and drug-related problems. For that, we restricted the initial demographically matched cohort to individuals with the follow-up longer than one year since the cohort entry (i.e., those who during the first year did not develop the outcome of interest or were not censored due to death, emigration from Sweden, end of study or – if unexposed – change of the exposure status). Thus, from the original 960,430 pairs, 59,825 (6.2%) and 62,687 (6.5%) matched pairs were excluded from the analyses of alcohol-related problems and drug-related problems, respectively. Cumulative dosage among BZDR-recipients was determined by summing the defined daily doses (DDD) dispensed during the first year after BZDR initiation and then categorized into quartiles: >0–<30, ≥30–<90, ≥90–<225, and ≥225 DDD. Then, we then calculated hazard ratios (HR) and 95% confidence intervals (CI) for any alcohol-related problems and, separately, drug-related problems comparing the BZDR-recipients belonging to cumulative doses categories to their matched comparators (i.e., 0 DDD). The follow-up started from the beginning of the second year after the cohort entry until the outcome of interest, death by non-outcome, emigration, change of the exposure status (for unexposed), and the study end, whichever occurred first. With the cumulative dosage from the first study year and the follow/up starting from the beginning of the second year the analysis was an analogy to intention to treat approach. We used full adjustment for modelling.

**Supplementary Table S1. ATC-codes retrieved from the Prescribed Drug Register for benzodiazepines, benzodiazepine-related Z-drugs, and other medications**

| **Medication** | **ATC-codes** | **Comments** |
| --- | --- | --- |
| **Benzodiazepines and benzodiazepine-related drugs retrieved from the PDR** | | |
| ***Benzodiazepine derivatives in antiepileptics*** | | |
| Clonazepam | N03AE01 |  |
| ***Benzodiazepine derivatives in anxiolytics*** | | |
| Diazepam | N05BA01 |  |
| Oxazepam | N05BA04 |  |
| Clorazepate | N05BA05 |  |
| Lorazepam | N05BA06 |  |
| Bromazepam | N05BA08 |  |
| Clobazam | N05BA09 |  |
| Alprazolam | N05BA12 |  |
| ***Benzodiazepine derivatives in hypnotics/sedatives*** | | |
| Nitrazepam | N05CD02 |  |
| Flunitrazepam | N05CD03 |  |
| Triazolam | N05CD05 |  |
| Midazolam | N05CD08 |  |
| ***Benzodiazepine-related drugs (Z-drugs)*** | | |
| Zopiclone | N05CF01 |  |
| Zolpidem | N05CF02 |  |
| Zaleplon | N05CF03 |  |
| **Other medications retrieved from the PDR, if recorded during 3 months prior to the cohort entry date** | | |
| Antidepressants | N06A |  |
| Psychostimulants (centrally acting sympathomimetics) | N06BA |  |
| Mood stabilisers | N03AF01, N03AF02, N03AG01, N03AX09, N03AN01 |  |
| (Non-BZD)-antiepileptics (i.e., excluding benzodiazepine derivatives) | N03 (except N03AE)^a^ and (except N03AF01, N03AF02, N03AG01, N03AX09, N03AN01)^b^ | ^a^ Excluded from non-BZD antiepileptics and used to select BZD derivatives in antiepileptics  ^b^ Excluded from non-BZD antiepileptics and used to select mood stabilizers |
| Antipsychotics | N05A |  |
| (Non-BZD)-anxiolytics (i.e., excluding benzodiazepine derivatives), and (non-BZDR)-hypnotics/sedatives (i.e., excluding benzodiazepine derivatives and benzodiazepine-related drugs) | N05B (except N05BA)^c^  N05C (except N05CD, N05CF)^d^ | ^c^ Excluded from non-BZD anxiolytics and used to select benzodiazepine derivatives in anxiolytics  ^d^ Excluded from non-BZDR hypnotics/sedatives and used to select benzodiazepine derivatives in hypnotics/sedatives (N05CD) and benzodiazepine-related drugs (N05CF) |
| Analgesics (non-opioids) | N02B, N02C |  |
| Opioids | N02A |  |

*Abbreviations*: ATC, Anatomical Therapeutic Chemical Classification System; BZD, benzodiazepines; BZDR, benzodiazepines and benzodiazepine-related Z-drugs when being mentioned together; PDR, Prescribed Drug Register

**Supplementary Table S2**. **ICD-codes, ATC-codes, and criminal offences codes for the ascertainment of alcohol-related problems and drug-related problems^a^**

| **Alcohol/drug-related problems** | **ICD-10 codes (1997-onwards),**  **ATC-codes (2005-onwards),**  **criminal offences codes (1995-onwards)** | **ICD-9 codes**  **(1987-1996)** | **ICD-8 codes (1969-1986)** |
| --- | --- | --- | --- |
| Alcohol use disorders or deaths | *ICD-10 codes:*  F10.1-F10.9 (Mental and behavioural disorders due to use of alcohol) | *ICD-9 codes:*  291 (Alcoholic psychosis),  303 (Alcoholism/ dependence), | *ICD-8 codes:*  291 (Alcoholic psychosis),  303 (Alcoholism/ dependence) |
| Medications used for alcohol dependence treatment^b^ | *ATC-codes*:  N07BB01 (disulfiram), N07BB03 (acamprosate), N07BB04 (naltrexone), N07BB05 (nalmefene) |  |  |
| Alcohol-related poisoning (non-fatal or fatal) | *ICD-10 codes:*  F10.0 (Acute intoxication by alcohol)  T51.0 (Toxic effect of ethanol),  X45 (Accidental poisoning by and exposure to alcohol) | *ICD-9 codes:*  305A (Alcohol misuse)  980A (Toxic effect of ethanol) | *ICD-8 codes:*  980,00 (Toxic effect of ethanol),  980,01 (Toxic effect of ethanol surrogates) |
| Alcohol-related criminal offences | *Criminal offences codes*:  3005 (Driving under the influence of only alcohol, or both alcohol and drugs; from the Law 1951:649, §4 and 4a),  3201 (Operating maritime vessel under the influence of alcohol or other drugs; from the Law 1994:1009, chapter 20, §4 and 5) |  |  |
| Drug-related disorders or deaths | *ICD-10 codes:*  F11.1-F11.9 (Mental and behavioural disorders due to use of opioids)  F12.1-F12.9 (...due to use of cannabinoids)  F13.1-F13.9 (...due to use of sedatives or hypnotics)  F14.1-F14.9 (...due to use of cocaine)  F15.1-F15.9 (...due to use of other stimulants)  F16.1-F16.9 (...due to use of hallucinogens)  F18.1-F18.9 (...due to use of volatile solvents)  F19.1-F19.9 (...due to multiple drug use of other psychoactive substances) | *ICD-9 codes:*  292 (Drug-induced psychoses)  304 (Drug dependence) | *ICD-8 codes:*  304 (Drug dependence) |
| Medications used in opioid use disorder treatment^b,c^ | *ATC-codes*:  N07BC01 (buprenorphine), N07BC02 (methadone), N07BC05 (levomethadone), and N07BC51 (buprenorphine, combinations) |  |  |
| Drug-related poisoning (non-fatal or fatal) | *ICD-10 codes:*  F11.0 (Acute intoxication due to use of opioids)  F12.0 (...due to use of cannabinoids)  F13.0 (...due to use of sedatives or hypnotics)  F14.0 (...due to use of cocaine)  F15.0 (...due to use of other stimulants)  F16.0 (...due to use of hallucinogens)  F18.0 (...due to use of volatile solvents)  F19.0 (...due to use of multiple drug use of other psychoactive substances)  T40 (Poisoning by narcotics and psychodysleptics),  T42 (Poisoning by antiepileptic, sedative-hypnotic and antiparkinsonism drugs)  X41 (Accidental poisoning by and exposure to antiepileptic, sedative-hypnotic, antiparkinsonism and psychotropic drugs, not elsewhere classified)  X42 (Accidental poisoning by and exposure to narcotics and psychodysleptics (hallucinogens), not elsewhere specified) | *ICD-9 codes:*  305X (Narcotic and medication misuse)  969 (Poisoning by psychotropic medications and narcotics)  969G (Poisoning by psychodysleptics) | *ICD-8 codes:*  971 (Poisoning by narcotics) |
| Drug-related criminal offenses | *Criminal offences codes*:  3070 (Driving under the influence of only drugs; from the Law 1951:649, paragraphs 4 and 4a),  5010 (Possession of drug only; from the Narcotic Drug Act 1968:64),  5011 (Use of drug only; from the Narcotic Drugs Act 1968:64),  5012 (Possession and use of drugs; from the Narcotic Drugs Act 1968:64) | | |

*Note*: The ICD-10 codes are included in the outcome definition, while together with ICD-8 and ICD-9, the codes are used for constructing variables ‘history of any alcohol use- or drug-related problems prior to the cohort entry’ (used as an additional exclusion criterion), and ‘maternal and paternal lifetime substance-related problems’ (used as a potential confounder).

^a^ ICD-codes are collected from the National Patient Register and the Cause of Death Register for non-fatal and fatal cases, respectively, ATC-codes are collected from the Prescribed Drug Register, and codes for criminal offences are collected from the Register of People Suspected of Offences.

^b^ Medications are selected according to the Swedish Pharmaceutical guidelines (www.fass.se)

^c^ Methadone or buprenorphine which are used in opioid use disorders therapy - namely N07BC01, N07BC02, N07BC05, and N07BC51 – were included in the outcome definition for the main analyses only. In the sensitivity analysis, these medications were excluded from outcome measures due to a possible misclassification between the use of methadone and buprenorphine for OUD therapy and pain therapy.

*Abbreviations*: ATC, Anatomical Therapeutic Chemical Classification System; BZDR, benzodiazepines and benzodiazepine-related Z-drugs; ICD, International Classification of Diseases.

**Supplementary Table S3. Psychiatric and somatic conditions collected from the National Patient Register, if recorded between 1997^a^ and the cohort entry date^b^**

| **Conditions** | **ICD-10 codes** |
| --- | --- |
| ***Psychiatric conditions, sleep disorders, and suicide attempt*** | |
| Schizophrenia, schizotypal, and delusional disorders, and psychotic disorders | F20, F21, F22, F23, F24, F25 (except F25.0), F28, F29 |
| Bipolar disorders | F25.0, F30, F31, F34.0 |
| Depressive disorders | F32, F33, F34 (except F34.0), F38, F39 |
| Anxiety disorders | F40, F41 |
| Obsessive-compulsive disorder | F42 |
| Reaction to severe stress and adjustment disorders | F43 |
| Dissociative, somatoform and other neurotic disorders | F44, F45, F48 |
| Mental retardation | F70-F79 |
| Autism spectrum disorders | F84.0, F84.1, F84.3, F84.5, F84.8, F84.9 |
| Attention Deficit / Hyperactivity Disorder (including medication for ADHD by ATC codes) | F90 and ATC-codes: N06BA01, N06BA02, N06BA04, N06BA09, N06BA12 |
| Disruptive behaviour disorders | F91 |
| Suicide attempt /self-injury (definite or undetermined intent) | X60–X84, Y10–Y34 |
| Nonorganic sleep disorders and insomnia (organic) | F51.0, G47.0 |
| ***Somatic conditions (chronic pain syndrome related conditions)*** | |
| Back and neck pain | M4, M5 |
| Headache | G43, G44, R51 |
| Arthritis, osteoarthritis, and joint pain | M0, M1, M2, R26 |
| Temporomandibular joint disorders | K07.6 |
| Fibromyalgia | M79.7 |
| Other musculoskeletal/connective tissue pain | M3, M6, M7, M8, M9 |
| Gastro-esophageal reflux disease | K21 |
| Irritable bowel syndrome | K58 |
| Chronic fatigue syndrome | G93.3 |
| Interstitial cystitis | N30.1 |
| Sexual pain | F52.5, F52.6, N94, N48.3 |
| ***Lifetime diagnosis of epilepsy used as the exclusion criterion for study population*** | |
| Epilepsy, status epilepticus | G40, G41 |

^a^ Introduction of the International Classification of Diseases, Tenth Revision (ICD-10) in Sweden

^b^ ‘Cohort entry’ refers to the date when the first prescription of any benzodiazepines or related Z-drugs (BZDR) was dispensed to the BZDR-recipient. The same date is assigned as the cohort entry to matched unexposed individual (in the demographically matched cohort) and to unexposed co-twin (in the co-twin control cohort)

*Abbreviations*: ADHD, Attention Deficit / Hyperactivity Disorder; ATC, Anatomical Therapeutic Chemical Classification System; ICD, International Classification of Diseases

**Supplementary Table S4. Characteristics of incident BZDR-recipients and unexposed individuals included in the demographically matched cohort and co-twin control cohort. Values are numbers (percentages) unless specified otherwise**

| **Characteristics** | **Demographically matched cohort** | | **Co-twin control cohort** | |
| --- | --- | --- | --- | --- |
|  | **BZDR-recipients**  **(n=960,430)** | **Matched unexposed (n=960,430)** | **BZDR-recipients**  **(n=12,048)** | **Unexposed co-twins (n=12,579)** |
| Women | 576,679 (60.0) | 576,679 (60.0) | 7356 (61.1) | 6535 (52.0) |
| Men | 383,751 (40.0) | 383,751 (40.0) | 4692 (38.9) | 6081 (48.0) |
| **Age at the cohort entry, years,**  **median (IQR)** | 51 (37-65) | 51 (37-65) | 50 (34-63) | 50 (34-63) |
| **Age at the cohort entry, years** |  |  |  |  |
| 10-17 | 4867 (0.5) | 4867 (0.5) | 107 (0.9) | 118 (0.9) |
| 18-29 | 134,098 (14.0) | 134,098 (14.0) | 2075 (17.2) | 2184 (17.4) |
| 30-64 | 570,680 (59.4) | 570,680 (59.4) | 7151 (59.4) | 7446 (59.2) |
| ≥65 | 250,785 (26.1) | 250,785 (26.1) | 2715 (22.5) | 2831 (22.5) |
| **Cohort entry year** |  |  |  |  |
| 2007-2009 | 282,969 (29.5) | 282,969 (29.5) | 3817 (31.7) | 4018 (31.9) |
| 2010-2012 | 235,751 (24.5) | 235,751 (24.5) | 3048 (25.3) | 3172 (25.2) |
| 2013-2015 | 220,133 (22.9) | 220,133 (22.9) | 2615 (21.7) | 2716 (21.6) |
| 2016-2019 | 221,577 (23.1) | 221,577 (23.1) | 2568 (21.3) | 2673 (21.2) |
| **Any psychiatric condition^a^** | 195,249 (20.3) | 81,402 (8.5) | 2460 (20.4) | 1110 (8.8) |
| **Chronic pain condition^b^** | 428,758 (44.6) | 331,915 (34.6) | 5076 (42.1) | 4139 (32.9) |
| **Any co-medication^c^** | 461,726 (48.1) | 94,827 (9.9) | 5935 (49.3) | 1332 (10.6) |
| **Maternal substance-related problems^c^** | 37,676 (3.9) | 26,420 (2.8) | 395 (3.3) | 407 (3.2) |
| **Paternal substance-related problems^c^** | 85,008 (8.9) | 67,619 (7.0) | 940 (7.8) | 982 (7.8) |
| **Household disposable income at the cohort entry^d^** |  |  |  |  |
| Lowest tertile | 290,296 (30.2) | 283,735 (29.5) | 3599 (29.9) | 3651 (29.0) |
| 2^nd^ tertile | 288,997 (30.1) | 285,512 (29.7) | 3639 (30.2) | 3609 (28.7) |
| Highest tertile | 281,615 (29.3) | 293,047 (30.5) | 3397 (28.2) | 3854 (30.6) |
| Unknown | 99,522 (10.4) | 98,136 (10.2) | 1413 (11.7) | 1465 (11.6) |
| **Follow-up duration,** **years, median (IQR)** |  |  |  |  |
| Alcohol-related problems | 7.6 (4.2-10.9) | 6.8 (3.6-10.2) | 7.9 (4.3-11.2) | 7.0 (3.8-10.5) |
| Drug-related problems | 7.7 (4.3-11.0) | 6.8 (3.7-10.2) | 7.9 (4.3-11.2) | 7.1 (3.9-10.6) |

^a^ Includes psychiatric disorders (schizophrenia, schizotypal, delusional, and psychotic disorders, bipolar disorders, depressive disorders, anxiety disorders, obsessive-compulsive disorder, reaction to severe stress and adjustment disorders, dissociative, somatoform and other neurotic disorders, mental retardation, autism spectrum disorders, attention deficit/hyperactivity disorder, disruptive behaviour disorders), suicide attempt/self-injury, nonorganic sleep disorders and organic insomnia, if recorded in the National Patient Register between 1997 (start of ICD-10) and the cohort entry date

^b^ Includes conditions related to chronic pain syndrome (back and neck pain, headache, arthritis, osteoarthritis, and joint pain, temporomandibular joint disorders, fibromyalgia, other musculoskeletal/connective tissue pain, gastro-esophageal reflux disease, irritable bowel syndrome, chronic fatigue syndrome, interstitial cystitis, and sexual pain), if recorded in the National Patient Register between 1997 (start of ICD-10) and the cohort entry date

^c^ Includes dispensation records of antidepressants, centrally acting sympathomimetics, mood stabilisers, non-BZD-antiepileptics, antipsychotics, non-BZD-anxiolytics, non-BZDR-hypnotics/sedatives, non-opioid analgesics, and opioids, if dispensed within 3 months before the cohort entry, according to the Prescribed Drug Register

^d^ Missing data on income were marked as unknown and included in the models as nominal variable

*Abbreviation*: BZD, benzodiazepines; BZDR, benzodiazepines and related Z-drugs (if mentioned together); ICD, International Classification of Diseases; IQR, interquartile range

**Supplementary Table S5**. **Associations of incident BZDR use with the risk of developing alcohol-related and drug-related problems subdivided by type of incident outcome events in 960,430 BZDR-recipients and 960,430 matched unexposed individuals in the demographically matched cohort**

|  | **No. of events, n (%)** | | **Crude incidence rate (95% CIs), per 1000 person-years** | | **Minimally-adjusted model^a^** | **Adjusted model^b^** | **Fully-adjusted model^c^** |
| --- | --- | --- | --- | --- | --- | --- | --- |
|  | **BZDR-recipients** | **Matched**  **unexposed** | **BZDR-recipients** | **Matched**  **unexposed** | **HR (95% CI)** | **HR (95% CI)** | **HR (95% CI)** |
| **Any alcohol-related problems** | | |  |  |  |  |  |
| Alcohol use disorders^d^ | 32,926 (3.43) | 13,265 (1.38) | 4.56 (4.51-4.61) | 1.99 (1.96-2.02) | 2.35 (2.30-2.40) | 1.75 (1.71-1.79) | 1.71 (1.67-1.75) |
| Poisoning | 562 (0.06) | 238 (0.02) | 0.08 (0.07-0.08) | 0.04 (0.03-0.04) | 2.23 (1.91-2.59) | 1.59 (1.33-1.89) | 1.54 (1.29-1.83) |
| Offences | 6623 (0.69) | 4862 (0.51) | 0.92 (0.89-0.94) | 0.73 (0.71-0.75) | 1.31 (1.26-1.36) | 1.18 (1.13-1.23) | 1.15 (1.11-1.20) |
| Deaths | 862 (0.09) | 568 (0.06) | 0.12 (0.11-0.13) | 0.09 (0.08-0.09) | 1.42 (1.27-1.58) | 1.24 (1.10-1.39) | 1.21 (1.08-1.37) |
| **Any drug-related problems** | | |  |  |  |  |  |
| Drug use disorders^e^ | 17,040 (1.77) | 3451 (0.36) | 2.34 (2.30-2.38) | 0.51 (0.50-0.53) | 4.70 (4.53-4.88) | 2.54 (2.44-2.65_ | 2.46 (2.36-2.56) |
| Poisoning | 2369 (0.25) | 376 (0.04) | 0.33 (0.31-0.34) | 0.06 (0.05-0.06) | 6.05 (5.43-6.75) | 3.42 (3.04-3.85) | 3.37 (2.99-3.79) |
| Offences | 10,733 (1.12) | 4465 (0.46) | 1.47 (1.45-1.50) | 0.67 (0.65-0.69) | 2.38 (2.30-2.46) | 1.79 (1.72-1.87) | 1.75 (1.68-1.83) |
| Deaths | 732 (0.08) | 185 (0.02) | 0.10 (0.09-0.11) | 0.03 (0.02-0.03) | 3.70 (3.15-4.35) | 2.27 (1.90-2.72) | 2.17 (1.81-2.60) |
| **Additional outcome within drug-related problems** | | | |  |  |  |  |
| Sedatives/hypnotics use disorders (non-fatal and fatal)^f^ | 4212 (0.44) | 512 (0.05) | 0.58 (0.56-0.60) | 0.08 (0.07-0.08) | 7.81 (7.13-8.56) | 4.26 (3.85-4.70) | 4.10 (3.71-4.53) |

*Note*: Some individuals who were defined as having ‘any alcohol-related problems’ may have more than one type of the outcome event that occurred at the same day (multiple simultaneous outcomes appeared in 1% of BZDR-recipients and in <2% of references). Same refers to individuals with ‘any drug-related problems’ (multiple simultaneous outcomes appeared in 2% of BZDR-recipients and in <3% of references). As a result, those individuals may appear in more than one analysis of the specific type of outcome.

^a^ Conditioned on matching variables (birth year and month, sex, and country of residence at the cohort entry)

^b^ Additionally adjusted for calendar year at the cohort entry, disposable family income, history of any psychiatric conditions, chronic pain conditions, concomitant dispensations of other psychotropic, antiepileptic, and analgesic medications, if dispensed within 3 months before the cohort entry, and history of maternal and paternal substance-related problems

^c^ Additionally adjusted for the events of the ‘other’ outcome if it was recorded during the study follow-up (i.e., controlling for drug-related problems in the analysis of alcohol-related problems, if occurred during the follow-up, and visa verse)

^d^ Including dispensation of medication for alcohol dependence

^e^ Including dispensation of medication for opioid use disorders

^f^ Additional type of outcome event within drug-related problems created by collecting the corresponding records from the National Patient Register and the Cause of Death Register. Due to data availability and in order to avoid underpowered analysis, this covariate combines the records of non-fatal and fatal cases of mental and behavioural disorders due to use of sedatives or hypnotics (ICD-10 codes: F13.1-F13.9) and non-fatal and fatal cases of poisoning by use of sedatives or hypnotics (ICD-10 code: F13.0)

*Abbreviation*: BZDR, benzodiazepines and benzodiazepine-related Z-drugs; CI, confidence intervals; HR, hazard ratio

**Supplementary Table S6**. **Associations of incident BZDR use with the risk of developing alcohol-related and drug-related problems subdivided by type of incident outcome events among 960,430 BZDR-recipients and 960,430 matched unexposed individuals from the demographically matched cohort at different follow-up periods**

|  | **Years since cohort entry** | | | | | | | |
| --- | --- | --- | --- | --- | --- | --- | --- | --- |
|  | **Alcohol-related problems** | | | | **Drug-related problems** | | | |
|  | **1 year** | **3 years** | **5 years** | **10 years** | **1 year** | **3 years** | **5 years** | **10 years** |
| **Disorders *(alcohol use disorders or, correspondingly, drug use disorders)*^a^** | | | | | | | | |
| HR (95% CI)^b^ | 1.65  (1.59-1.71) | 1.54  (1.48-1.59) | 1.62  (1.57-1.67) | 1.83  (1.75-1.91) | 2.57  (2.42-2.73) | 2.14  (2.01-2.27) | 2.20  (2.09-2.31) | 2.38  (2.19-2.59) |
| Cumulative incidence^c^ (95% CI), % |  |  |  |  |  |  |  |  |
| BZDR-recipients | 0.65  (0.64-0.67) | 1.42  (1.40-1.44) | 2.03  (2.01-2.06) | 3.36  (3.32-3.41) | 0.34  (0.33-0.35) | 0.72  (0.70-0.73) | 0.99  (0.98-1.01) | 1.60  (1.57-1.62) |
| Unexposed individuals | 0.32  (0.31-0.33) | 0.80  (0.78-0.82) | 1.16  (1.14-1.19) | 1.84  (1.80-1.87) | 0.11  (0.10-0.11) | 0.26  (0.25-0.28) | 0.39  (0.37-0.40) | 0.62  (0.60-0.64) |
| Difference^d^ | 0.33  (0.31-0.35) | 0.62  (0.59-0.65) | 0.87  (0.83-0.91) | 1.53  (1.47-1.58) | 0.24  (0.22-0.25) | 0.45  (0.43-0.47) | 0.61  (0.58-0.63) | 0.98  (0.94-1.02) |
| **Poisoning *(by a corresponding substance)*** | | | | | | | | |
| HR (95% CI)^b^ | 1.31  (0.96-1.79) | 1.52 (1.14-2.04) | 1.56  (1.23-1.99) | 1.47  (1.11-1.95) | 3.20  (2.71-3.78) | 2.35  (1.98-2.80) | 2.36  (2.03-2.76) | 2.41  (1.85-3.14) |
| Cumulative incidence^c^ (95% CI), % |  |  |  |  |  |  |  |  |
| BZDR-recipients | 0.01  (0.01-0.01) | 0.02 (0.02-0.02) | 0.03  (0.03-0.04) | 0.07  (0.06-0.08) | 0.08  (0.07-0.08) | 0.13  (0.12-0.14) | 0.16  (0.15-0.17) | 0.20  (0.19-0.21) |
| Unexposed individuals | 0.01  (0.00-0.01) | 0.01 (0.01-0.02) | 0.02  (0.02-0.02) | 0.04  (0.03-0.05) | 0.01  (0.01-0.02) | 0.03  (0.03-0.04) | 0.04  (0.04-0.05) | 0.06  (0.05-0.06) |
| Difference^d^ | 0.01  (0.00-0.01) | 0.01 (0.00-0.01) | 0.01  (0.01-0.02) | 0.03  (0.02-0.04) | 0.07  (0.06-0.07) | 0.10  (0.09-0.11) | 0.12  (0.11-0.13) | 0.14  (0.13-0.15) |
| **Offences *(related to use of a corresponding substance)*** | | | | | | | | |
| HR (95% CI)^b^ | 1.08  (1.01-1.15) | 1.13  (1.06-1.20) | 1.18  (1.11-1.24) | 1.24  (1.14-1.35) | 1.72  (1.62-1.83) | 1.58  (1.48-1.68) | 1.74 (1.65-1.83) | 2.02  (1.87-2.19) |
| Cumulative incidence^c^ (95% CI), % |  |  |  |  |  |  |  |  |
| BZDR-recipients | 0.12  (0.11-0.12) | 0.30  (0.29-0.31) | 0.45  (0.44-0.46) | 0.74  (0.72-0.76) | 0.21  (0.20-0.22) | 0.50  (0.49-0.52) | 0.71 (0.70-0.73) | 1.20  (1.17-1.22) |
| Unexposed individuals | 0.10  (0.10-0.11) | 0.26  (0.25-0.27) | 0.38  (0.37-0.39) | 0.58  (0.57-0.60) | 0.11  (0.11-0.12) | 0.29  (0.28-0.31) | 0.42 (0.40-0.43) | 0.66  (0.64-0.68) |
| Difference^d^ | 0.01  (0.01-0.02) | 0.04  (0.02-0.05) | 0.07  (0.05-0.09) | 0.16  (0.13-0.18) | 0.09  (0.08-0.11) | 0.21  (0.19-0.23) | 0.29 (0.27-0.32) | 0.54  (0.50-0.57) |
| **Deaths *(due to use of a corresponding substance)*** | | | | | | | | |
| HR (95% CI)^b^ | 1.10  (0.88-1.38) | 1.09  (0.92-1.29) | 1.17  (0.97-1.41) | 1.40  (1.16-1.69) | 1.46  (1.08-1.97) | 2.14  (1.65-2.77) | 2.71  (2.01-3.65) | 2.67  (1.95-3.66) |
| Cumulative incidence^c^ (95% CI), % |  |  |  |  |  |  |  |  |
| BZDR-recipients | 0.01  (0.01-0.01) | 0.03  (0.03-0.03) | 0.05  (0.05-0.06) | 0.12  (0.11-0.13) | 0.01  (0.00-0.01) | 0.02  (0.02-0.02) | 0.04  (0.03-0.04) | 0.08  (0.08-0.09) |
| Unexposed individuals | 0.01  (0.01-0.01) | 0.03  (0.02-0.03) | 0.04  (0.04-0.05) | 0.08  (0.07-0.09) | 0.00  (0.00-0.01) | 0.01  (0.01-0.02) | 0.02  (0.02-0.02) | 0.03  (0.03-0.04) |
| Difference^d^ | 0.00  (0.00-0.00) | 0.00  (0.00-0.01) | 0.01  (0.00-0.02) | 0.04  (0.03-0.05) | 0.00  (0.00-0.00) | 0.01  (0.00-0.01) | 0.02  (0.01-0.02) | 0.05  (0.04-0.06) |

*Note*: Cumulative incidence measures in BZDR-recipients and unexposed individuals represent absolute risks of outcomes in these groups at each specific follow-up time, and cumulative incidence differences represent the corresponding absolute risk differences.

^a^ For alcohol-related problems (left part of the table): diagnoses of alcohol use disorders also include dispensation records of medication for alcohol dependence; for drug-related problems (right part of the table): diagnoses of drug use disorders alco include dispensation records of medication for opioid use disorder

^b^ All reported hazard ratios represent the results of fully-adjusted model for the demographically matched cohort

^c^ All reported cumulative incidences are standardised, i.e., controlled for the covariates which were included in fully-adjusted model for the demographically matched cohort

^d^ Cumulative incidences difference (in percentage) shows the excess outcome cases per 100 individuals in exposed compared to unexposed

*Abbreviation*: BZDR, benzodiazepines and benzodiazepine-related Z-drugs; CI, confidence intervals; HR, hazard ratio

**Supplementary Table S7**. **Associations of incident BZDR use with the risk of developing any alcohol-related problems stratified by study characteristics in 960,430 BZDR-recipients and 960,430 matched unexposed individuals in the demographically matched cohort**

|  | **No. of events, n (%)** | | **Crude incidence rate (95% CIs), per 1000 person years** | | **Fully-adjusted model^a^** |
| --- | --- | --- | --- | --- | --- |
|  | **BZDR-recipients** | **Matched**  **unexposed** | **BZDR-recipients** | **Matched**  **unexposed** | **HR (95 % CI)** |
| **Sex** |  |  |  |  |  |
| Women | 17,512 (3.04) | 7291 (1.26) | 3.90 (3.84-3.96) | 1.83 (1.79-1.87) | 1.51 (1.46-1.55) |
| Men | 22,963 (5.98) | 11,290 (2.94) | 8.39 (8.28-8.50) | 4.21 (4.14-4.29) | 1.62 (1.58-1.66) |
| **Age at cohort entry, years** |  |  |  |  |  |
| 10-29 | 9579 (6.89) | 3203 (2.30) | 8.94 (8.76-9.12) | 3.16 (3.05-3.27) | 1.79 (1.71-1.88) |
| 30-64 | 26,731 (4.68) | 12,427 (2.18) | 5.80 (5.74-5.87) | 2.94 (2.89-2.99) | 1.59 (1.55-1.62) |
| ≥65 | 4165 (1.66) | 2951 (1.18) | 2.69 (2.61-2.77) | 2.08 (2.00-2.16) | 1.25 (1.19-1.32) |
| **Cohort entry year** |  |  |  |  |  |
| 2007-2009 | 17,567 (6.21) | 9112 (3.22) | 5.50 (5.42-5.58) | 3.28 (3.22-3.35) | 1.35 (1.31-1.39) |
| 2010-2012 | 11,323 (4.80) | 5010 (2.13) | 5.55 (5.45-5.65) | 2.61 (2.54-2.69) | 1.62 (1.56-1.68) |
| 2013-2015 | 4109 (1.85) | 1525 (0.69) | 6.21 (6.02-6.40) | 2.29 (2.17-2.4) | 1.90 (1.81-1.99) |
| 2016-2019 | 7476 (3.40) | 2934 (1.33) | 5.63 (5.50-5.76) | 2.25 (2.17-2.33) | 2.13 (1.99-2.27) |
| **Any psychiatric condition^b^** |  |  |  |  |  |
| Yes | 12,851 (6.58) | 2795 (3.43) | 9.10 (8.94-9.26) | 6.11 (5.89-6.34) | 1.22 (1.17-1.28) |
| No | 27,624 (3.61) | 15,786 (1.80) | 4.75 (4.70-4.81) | 2.54 (2.50-2.58) | 1.63 (1.59-1.66) |
| **Chronic pain condition^c^** |  |  |  |  |  |
| Yes | 15,870 (3.70) | 5936 (1.79) | 5.33 (5.25-5.42) | 2.95 (2.88-3.03) | 1.45 (1.40-1.50) |
| No | 24,605 (4.63) | 12,645 (2.01) | 5.79 (5.72-5.86) | 2.72 (2.67-2.76) | 1.61 (1.57-1.65) |
| **Any co-medication^d^** |  |  |  |  |  |
| Yes | 22,674 (4.91) | 2367 (2.50) | 6.84 (6.75-6.93) | 4.38 (4.20-4.56) | 1.19 (1.13-1.24) |
| No | 17,801 (3.57) | 16,214 (1.87) | 4.56 (4.49-4.62) | 2.65 (2.61-2.69) | 1.61 (1.57-1.64) |
| **BZDR at initiation^e^** |  |  |  |  |  |
| BZD | 15,908 (4.40) | 6931 (1.92) | 5.93 (5.83-6.02) | 2.76 (2.7-2.83) | 1.59 (1.55-1.63) |
| Z-drugs | 24,567 (4.10) | 11,650 (1.95) | 5.41 (5.34-5.48) | 2.80 (2.75-2.85) | 1.55 (1.52-1.63) |

*Note*: For all covariates, p-values for interaction between subgroup HRs are *p*<0.001. For BZDR at initiation, p-value from Wald test is *p*=0.013.

^a^ All reported hazard ratios represent the results of fully-adjusted model for the demographically matched cohort

^b^ Includes psychiatric disorders (schizophrenia, schizotypal, and delusional disorders, and psychotic disorders, bipolar disorders, depressive disorders, anxiety disorders, obsessive-compulsive disorder, reaction to severe stress and adjustment disorders, dissociative, somatoform and other neurotic disorders, mental retardation, autism spectrum disorders, attention deficit/hyperactivity disorder, disruptive behaviour disorders), suicide attempt/self-injury, nonorganic sleep disorders and organic insomnia, if recorded in the National Patient Register between 1997 (the start of ICD-10) and the cohort entry date

^c^ Includes conditions related to chronic pain syndrome (back and neck pain, headache, arthritis, osteoarthritis, and joint pain, temporomandibular joint disorders, fibromyalgia, other musculoskeletal/connective tissue pain, gastro-esophageal reflux disease, irritable bowel syndrome, chronic fatigue syndrome, interstitial cystitis, and sexual pain), if recorded in the National Patient Register between 1997 (the start of ICD-10) and the cohort entry date

^d^ Includes dispensation records of antidepressants, centrally acting sympathomimetics, mood stabilisers, non-BZD-antiepileptics, antipsychotics, non-BZD-anxiolytics, non-BZDR-hypnotics/sedatives, non-opioid analgesics, and opioids, if dispensed within 3 months before the cohort entry, according to the Prescribed Drug Register

^e^ Type of medication (either any BZDs [N03AE, N05BA, or N05CD] or Z-drugs [N05CF]), which was collected at the first BZDR dispensation of in 2007-2019

*Abbreviation*: BZD, benzodiazepines; BZDR, benzodiazepines and benzodiazepine-related Z-drugs; ICD, International Classification of Diseases; IQR, interquartile range

**Supplementary Table S8**. **Cumulative BZDR dosage (in DDDs) during the first year after BZDR initiation and the risk of developing alcohol-related problems (in 900,605 BZDR-recipients versus 900,605 matched unexposed individuals) and drug-related problems (in 897,743 BZDR-recipients versus 897,743 matched unexposed individuals), with the follow-up starting from the second year after BZDR initiation**

| **Cumulative BZDR dosage in the first year**  **(in the defined daily dose)** | **No. of events, n (%)** | **Crude incidence rate (95% CIs), per 1000 person years** | **Fully-adjusted model,**  **HR (95% CI)** |
| --- | --- | --- | --- |
| **Alcohol-related problems** |  |  |  |
| 0 (ref – matched unexposed) | 6225 (0.69) | 1.11 (1.08-1.13) | 1.00 |
| >0 - <30 | 8721 (1.87) | 2.79 (2.73-2.85) | 2.23 (2.14-2.33) |
| ≥30 - <90 | 6166 (2.37) | 3.42 (3.33-3.50) | 3.07 (2.90-3.25) |
| ≥90 - <225 | 3945 (3.28) | 4.84 (4.69-50) | 4.94 (4.53-5.38) |
| ≥225 | 3221 (5.90) | 9.63 (9.30-9.97) | 10.64 (9.31-12.15) |
| **Drug-related problems** |  |  |  |
| 0 (ref - matched unexposed) | 14.551 (1.62) | 2.61 (2.56-2.65) | 1.00 |
| >0 - <30 | 13,728 (2.96) | 4.45 (4.38-4.52) | 1.65 (1.60-1.70) |
| ≥30 - <90 | 9151 (3.53) | 5.13 (5.03-5.24) | 1.89 (1.82-1.96) |
| ≥90 - <225 | 5098 (4.24) | 6.33 (6.15-6.50) | 2.32 (2.19-2.45) |
| ≥225 | 2945 (5.37) | 8.78 (8.46-9.09) | 2.95 (2.72-3.21) |

*Note*: In this analysis, only individuals with the follow-up longer than one year since the cohort entry were included (i.e., those who during the first year did not develop the outcome of interest or were not censored due to death, emigration from Sweden, end of study or – if unexposed – change of the exposure status). Therefore, from the original 960,430 pairs in the demographically matched cohort, 59,825 (6.2%) and 62,687 (6.5%) matched pairs were excluded from the analyses of alcohol-related problems and drug-related problems, respectively.

*Abbreviation*: BZDR, benzodiazepines and benzodiazepine-related Z-drugs; CI, confidence intervals; DDD, defined daily dose; HR, hazard ratio

**Supplementary Table S9**. **Associations of incident BZDR use with the risk of developing any drug-related problems stratified by study characteristics in 960,430 BZDR-recipients and 960,430 matched unexposed individuals in the demographically matched cohort**

|  | **No. of events, n (%)** | | **Crude incidence rate (95% CIs), per 1000 person years** | | **Fully-adjusted model^a^** |
| --- | --- | --- | --- | --- | --- |
|  | **BZDR-recipients** | **Matched unexposed** | **BZDR-recipients** | **Matched unexposed** | **HR (95% CI)** |
| **Sex** |  |  |  |  |  |
| Women | 14,809 (2.57) | 3520 (0.61) | 3.29 (3.24-3.34) | 0.88 (0.85-0.91) | 2.12 (2.04-2.21) |
| Men | 15,419 (4.02) | 4707 (1.23) | 5.55 (5.46-5.64) | 1.74 (1.69-1.79) | 2.12 (2.04-2.20) |
| **Age at cohort entry, years** |  |  |  |  |  |
| 10-29 | 14,717 (10.59) | 4270 (3.07) | 14.11 (13.88-14.34) | 4.22 (4.10-4.35) | 2.24 (2.15-2.33) |
| 30-64 | 13,110 (2.30) | 3166 (0.55) | 2.80 (2.75-2.85) | 0.74 (0.72-0.77) | 2.17 (2.08-2.26) |
| ≥65 | 2401 (0.96) | 791 (0.32) | 1.54 (1.48-1.60) | 0.55 (0.52-0.59) | 1.95 (1.79-2.13) |
| **Cohort entry year** |  |  |  |  |  |
| 2007-2009 | 11,920 (4.21) | 3534 (1.25) | 3.69 (3.62-3.75) | 1.26 (1.22-1.30) | 1.87 (1.80-1.96) |
| 2010-2012 | 8431 (3.58) | 2357 (1.00) | 4.10 (4.02-4.19) | 1.22 (1.17-1.27) | 2.03 (1.92-2.14) |
| 2013-2015 | 6134 (2.79) | 1518 (0.69) | 4.60 (4.49-4.72) | 1.16 (1.10-1.22) | 2.45 (2.29-2.61) |
| 2016-2019 | 3743 (1.69) | 818 (0.37) | 5.65 (5.47-5.83) | 1.23 (1.14-1.31) | 2.85 (2.61-3.10) |
| **Any psychiatric disorder^b^** |  |  |  |  |  |
| Yes | 12,766 (6.54) | 1743 (2.14) | 9.03 (8.88-9.19) | 3.78 (3.60-3.96) | 1.57 (1.49-1.66) |
| No | 17,462 (2.28) | 6484 (0.74) | 2.98 (2.93-3.02) | 1.04 (1.01-1.06) | 2.21 (2.14-2.29) |
| **Chronic pain condition^c^** |  |  |  |  |  |
| Yes | 12,406 (2.89) | 2807 (0.85) | 4.14 (4.07-4.21) | 1.39 (1.34-1.44) | 1.80 (1.72-1.88) |
| No | 17,822 (3.35) | 5420 (0.86) | 4.16 (4.10-4.22) | 1.16 (1.13-1.19) | 2.34 (2.25-2.42) |
| **Any co-medication^d^** |  |  |  |  |  |
| Yes | 20,136 (4.36) | 1341 (1.41) | 6.04 (5.96-6.12) | 2.46 (2.33-2.6) | 1.60 (1.51-1.69) |
| No | 10,092 (2.02) | 6886 (0.80) | 2.56 (2.51-2.61) | 1.12 (1.09-1.14) | 2.22 (2.15-2.29) |
| **BZDR at initiation^e^** |  |  |  |  |  |
| BZD | 11,797 (3.26) | 3154 (0.87) | 4.35 (4.27-4.43) | 1.25 (1.21-1.29) | 2.00 (1.93-2.06) |
| Z-drugs | 18,431 (3.08) | 5073 (0.85) | 4.03 (3.97-4.09) | 1.21 (1.18-1.25) | 2.17 (2.11-2.24) |

*Note*: For all covariates except sex, p-values for interaction between subgroup HRs are *p*<0.001. For BZDR at initiation, p-value from Wald test is *p*<0.001.

^a^ All reported hazard ratios represent the results of fully-adjusted model for the demographically matched cohort

^b^ Includes psychiatric disorders (schizophrenia, schizotypal, and delusional disorders, and psychotic disorders, bipolar disorders, depressive disorders, anxiety disorders, obsessive-compulsive disorder, reaction to severe stress and adjustment disorders, dissociative, somatoform and other neurotic disorders, mental retardation, autism spectrum disorders, attention deficit/hyperactivity disorder, disruptive behaviour disorders), suicide attempt/self-injury, nonorganic sleep disorders and organic insomnia, if recorded in the National Patient Register between 1997 (the start of ICD-10) and the cohort entry date

^c^ Includes conditions related to chronic pain syndrome (back and neck pain, headache, arthritis, osteoarthritis, and joint pain, temporomandibular joint disorders, fibromyalgia, other musculoskeletal/connective tissue pain, gastro-esophageal reflux disease, irritable bowel syndrome, chronic fatigue syndrome, interstitial cystitis, and sexual pain), if recorded in the National Patient Register between 1997 (the start of ICD-10) and the cohort entry date

^d^ Includes dispensation records of antidepressants, centrally acting sympathomimetics, mood stabilisers, non-BZD-antiepileptics, antipsychotics, non-BZD-anxiolytics, non-BZDR-hypnotics/sedatives, non-opioid analgesics, and opioids, if dispensed within 3 months before the cohort entry, according to the Prescribed Drug Register

^e^ Type of medication (either any BZDs [N03AE, N05BA, or N05CD] or Z-drugs [N05CF]), which was collected at the first BZDR dispensation in 2007-2019

*Abbreviation*: BZD, benzodiazepines; BZDR, benzodiazepines and benzodiazepine-related Z-drugs; ICD, International Classification of Diseases; IQR, interquartile range

**Supplementary Table S10**. **Sensitivity analysis within the demographically matched cohort using the new definition of drug-related outcomes (i.e., without dispensation records for opioid use disorders medication) in 960,430 BZDR-recipients and 960,430 matched unexposed individuals**

|  | **No. of events, n (%)** | | **Crude incidence rate (95% CIs), per 1000 person-years** | | **Minimally-adjusted model^a^** | **Adjusted**  **model^b^** | **Fully-adjusted model^c^** |
| --- | --- | --- | --- | --- | --- | --- | --- |
|  | **BZDR-recipients** | **Matched**  **unexposed** | **BZDR-recipients** | **Matched unexposed** | **HR (95% CI)** | **HR (95% CI)** | **HR (95% CI)** |
| Any drug-related problems | 26,816 (2.79) | 7126 (0.74) | 3.68 (3.64-3.72) | 1.06 (1.04-1.09) | 3.65 (3.56-3.75) | 2.25 (2.18-2.32) | 2.18 (2.11-2.24) |
| *By type of events^d^:* |  |  |  |  |  |  |  |
| Drug use disorders | 13,592 (1.42) | 2348 (0.24) | 1.86 (1.83-1.9) | 0.35 (0.34-0.36) | 5.55 (5.32-5.80) | 2.91 (2.77-3.06) | 2.79 (2.66-2.93) |
| Poisoning | 2388 (0.25) | 376 (0.04) | 0.33 (0.31-0.34) | 0.06 (0.05-0.06) | 6.09 (5.47-6.79) | 3.43 (3.05-3.86) | 3.38 (3.00-3.80) |
| Offences | 10,745 (1.12) | 4466 (0.47) | 1.47 (1.45-1.5) | 0.67 (0.65-0.69) | 2.38 (2.30-2.46) | 1.79 (1.72-1.87) | 1.75 (1.68-1.83) |
| Deaths | 737 (0.08) | 185 (0.02) | 0.10 (0.09-0.11) | 0.03 (0.02-0.03) | 3.72 (3.17-4.37) | 2.27 (1.89-2.71) | 2.17 (1.81-2.60) |

^a^ Conditioned on matching variables (birth year and month, sex, country of residence at the cohort entry)

^b^ Additionally adjusted for calendar year at the cohort entry, disposable family income, history of any psychiatric conditions, chronic pain conditions, concomitant dispensations of other psychotropic, antiepileptic, and analgesic medications, if dispensed within 3 months before the cohort entry, and history of maternal and paternal substance-related problems

^c^ Additionally adjusted for the events of any alcohol-related outcome if it was recorded during the study follow-up (i.e., controlling for drug-related problems in the analysis of alcohol-related problems and visa verse)

^d^ Some individuals who, by the new definition, were defined as having ‘any drug-related problems’ may have more than one type of the outcome event that occurred at the same day (multiple simultaneous outcomes appeared in 2% of BZDR-recipients and in less than 3% of references). As a result, those individuals may appear in more than one analysis of the specific type of outcome.

*Abbreviation*: BZDR, benzodiazepines and benzodiazepine-related Z-drugs; CI, confidence intervals; HR, hazard ratio

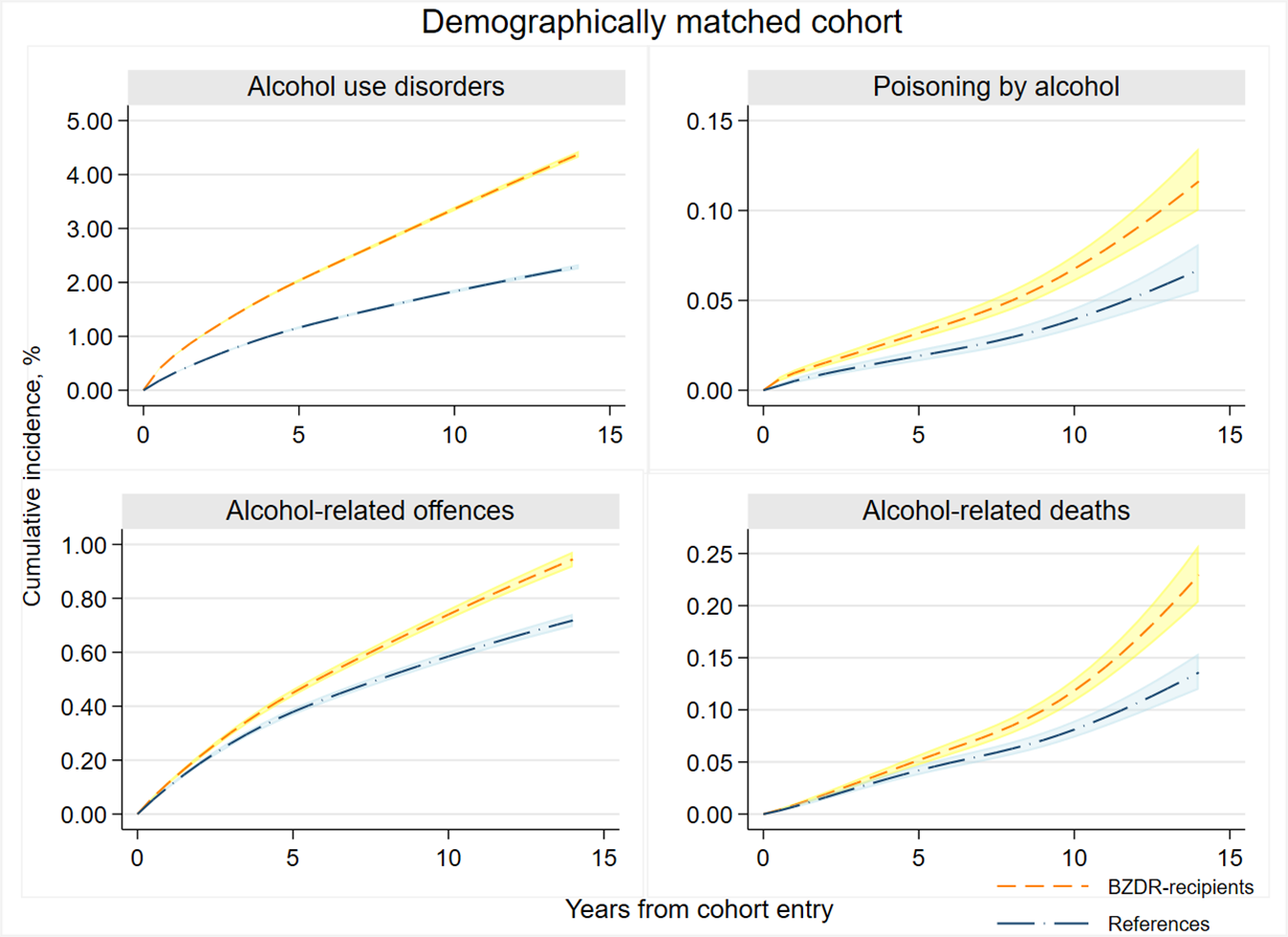

**Supplementary Figure S1**. **Standardized cumulative incidence and 95% CI of various types of incident outcome events within ‘any alcohol-related problems’ estimated as a function of time since the cohort entry among BZDR-recipients and their matched comparators (references) in the demographically matched cohort**

*Note*: Cumulative incidence measures are standardised, i.e., controlled for the covariates which were included in fully adjusted model for the demographically matched cohort, and estimated by the flexible parametric model. Shadowed areas denote 95% CI. The events indicated as ‘alcohol use disorders’ refer to the diagnoses and dispensations of medication for alcohol dependence.

*Abbreviations*: BZDR, benzodiazepine and benzodiazepine-related Z-drugs; CI, confidence intervals.

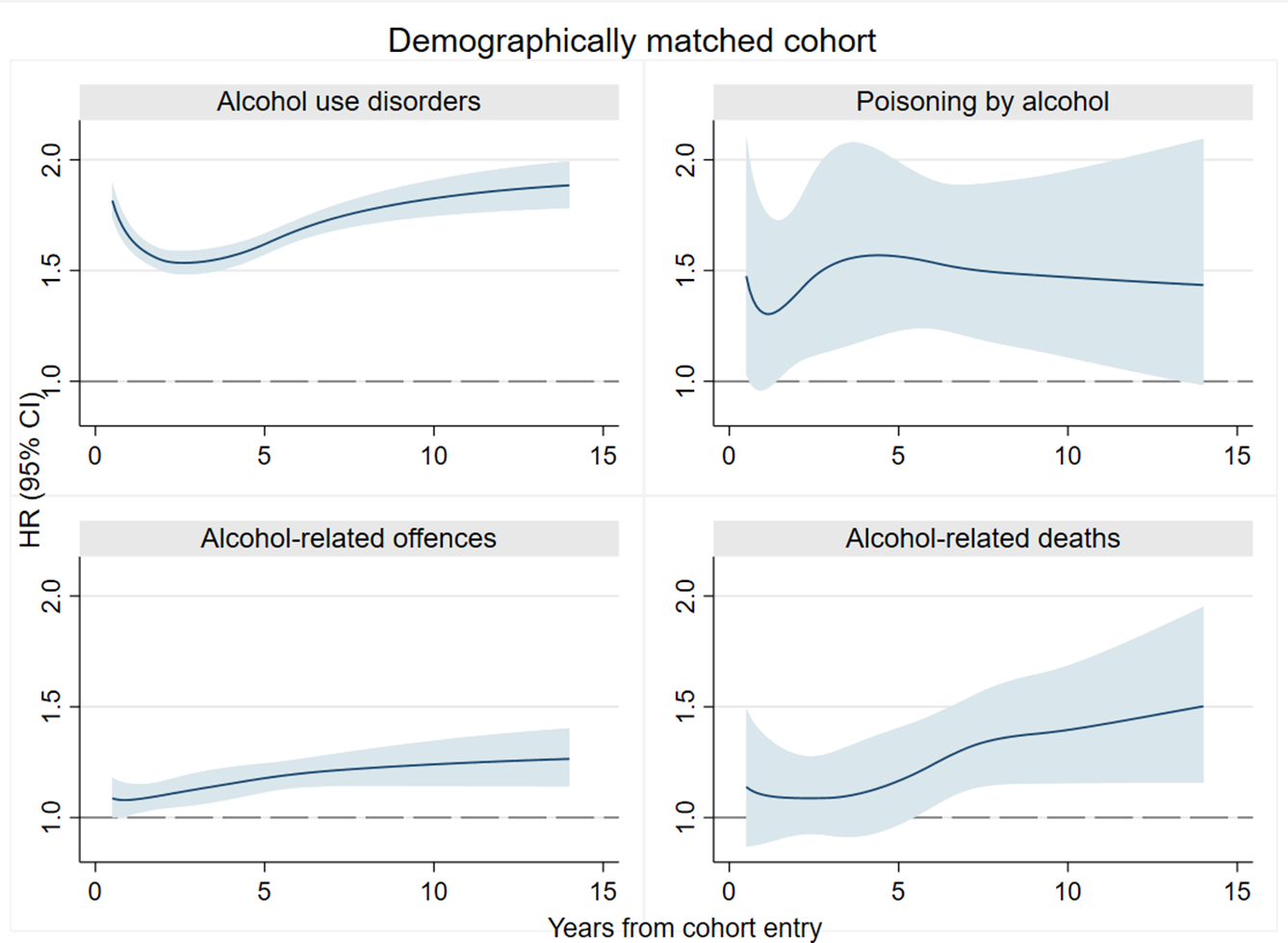

**Supplementary Figure S2**. **The risk of developing various types of incident outcome events within ‘any alcohol-related problems’ estimated as a function of time since the cohort entry in BZDR-recipients and their matched comparators (references) in the demographically matched cohort**

*Note*: All reported HRs represent the results from fully-adjusted model for the demographically matched cohort. Shadowed areas denote 95% CI. The events indicated as ‘alcohol use disorders’ refer to the diagnoses and dispensations of medication for alcohol dependence.

*Abbreviation*: BZDR, benzodiazepine and benzodiazepine-related Z-drugs; CI, confidence intervals; HR, hazard ratio

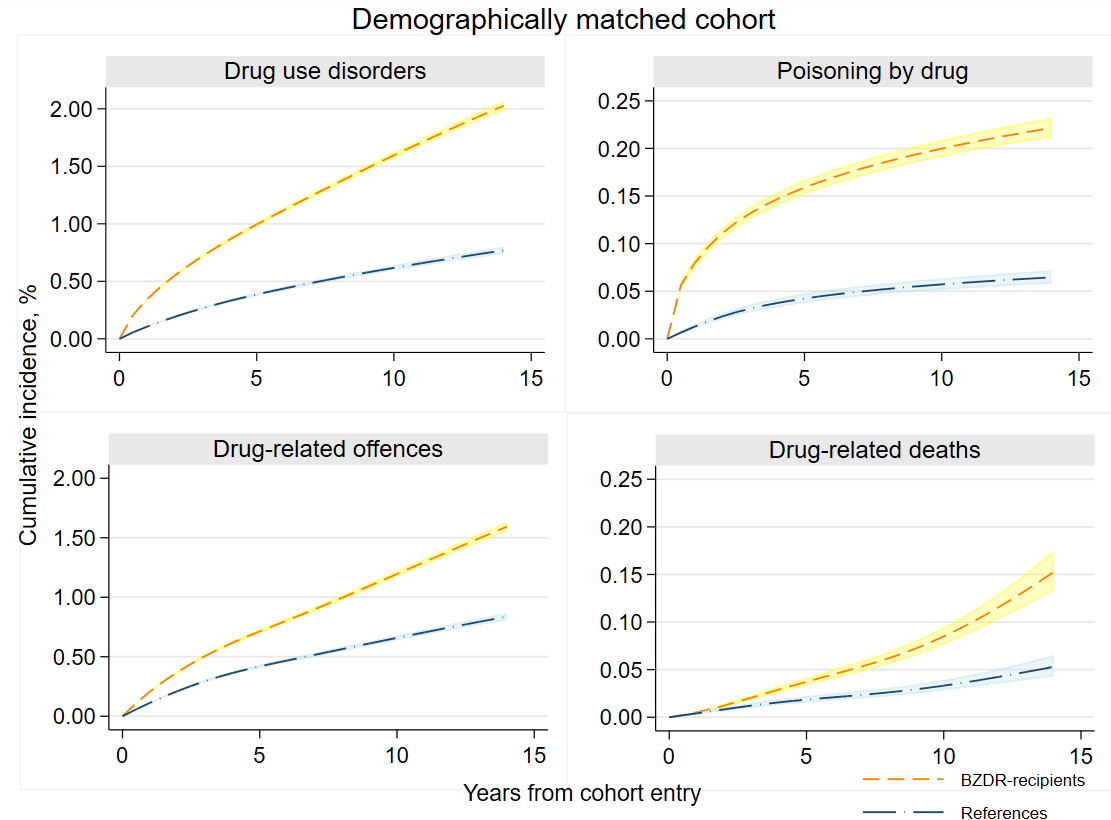

**Supplementary Figure S3**. **Standardized cumulative incidence and 95% CI of various types of incident outcome events within ‘any drug-related problems’ estimated as a function of time since the cohort entry among BZDR-recipients and their matched comparators (references) in the demographically matched cohort**

*Note*: Cumulative incidence measures are standardised, i.e., controlled for the covariates which were included in fully adjusted model for the demographically matched cohort, and estimated by the flexible parametric model. Shadowed areas denote 95% CI. The events indicated as ‘drug use disorders’ refer to the diagnoses and dispensations of medication for opioid use disorders.

*Abbreviations*: BZDR, benzodiazepine and benzodiazepine-related Z-drugs; CI, confidence intervals

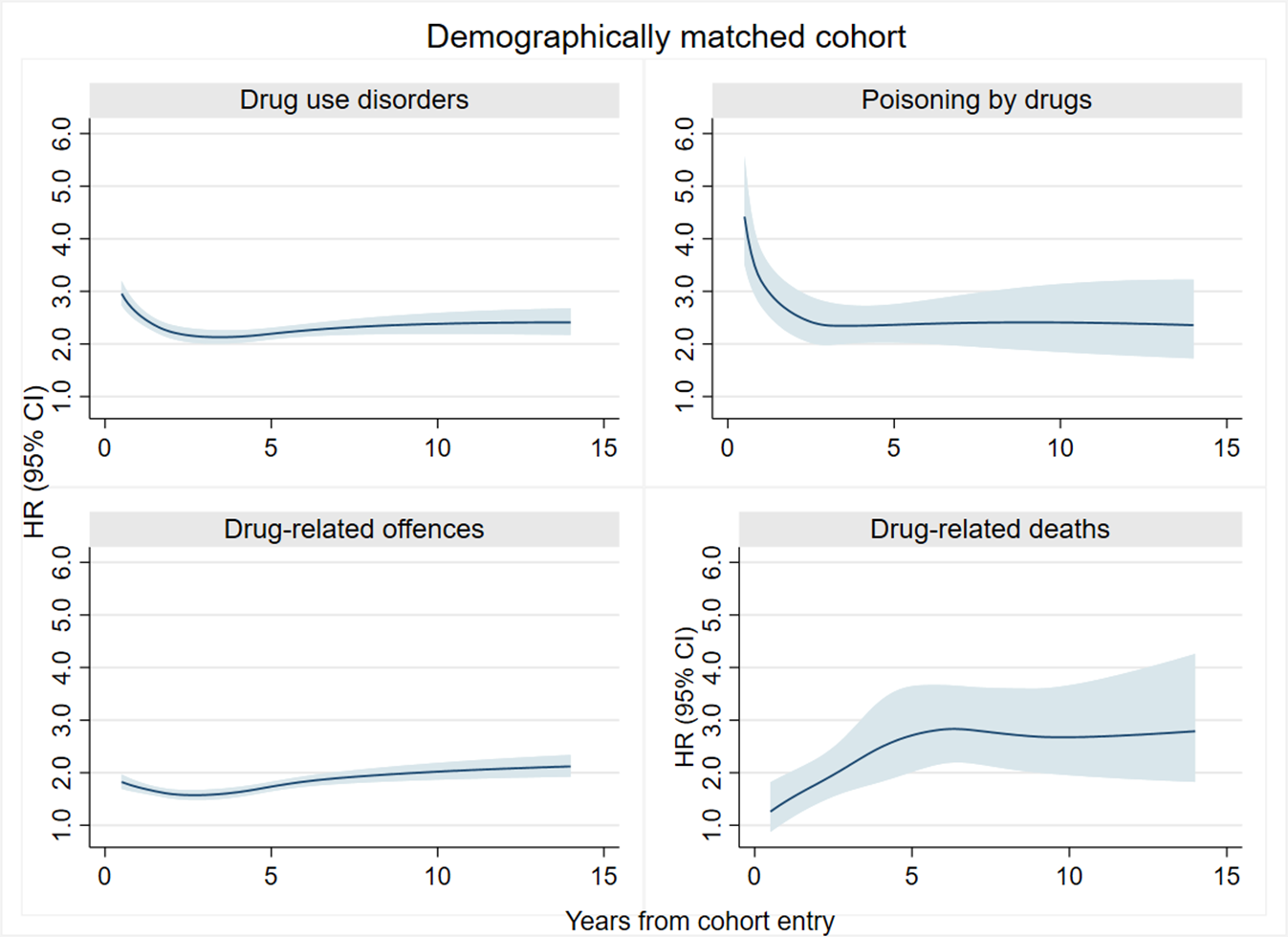

**Supplementary Figure S4**. **The risk of developing various types of incident outcome events as within ‘any drug-related problems’ estimated as a function of time since the cohort entry in BZDR-recipients and their matched comparators (references) in the demographically matched cohort**

*Note*: All reported HRs represent the results of fully-adjusted model for the demographically matched cohort. Shadowed areas denote 95% CI. The events indicated as ‘drug use disorders’ refer to the diagnoses and dispensations of medication for opioid use disorders.

*Abbreviation*: BZDR, benzodiazepine and benzodiazepine-related Z-drugs; CI, confidence intervals; HR, hazard ratio.
